# An elevated rate of whole-genome duplications associated with carcinogen exposure in Black cancer patients

**DOI:** 10.1101/2023.11.10.23298349

**Authors:** Leanne M. Brown, Ryan A. Hagenson, Tilen Koklič, Iztok Urbančič, Janez Strancar, Jason M. Sheltzer

## Abstract

In the United States, Black individuals have higher rates of cancer mortality than any other racial or ethnic group. The sources of these significant racial disparities are not fully understood, and may include social, environmental, and genetic factors that influence cancer onset, diagnosis, and treatment. Here, we examined genomic data from several large-scale cancer patient cohorts to search for racial associations in chromosome copy number alterations. We found that tumors from self-reported Black patients were significantly more likely to exhibit whole-genome duplications (WGDs), a genomic event that enhances metastasis and aggressive disease, compared to tumors from self-reported white patients. Among patients with WGD-positive cancers, there was no significant difference in survival between self-reported Black and white patients, suggesting that the increased incidence of WGD events could contribute to the disparities in patient outcome. We further demonstrate that combustion byproducts are capable of driving genome-duplication events in cell culture, and cancers from self-reported Black patients exhibit mutational patterns consistent with increased exposure to these carcinogens. In total, these findings identify a class of genomic alterations that are associated with environmental exposures and that may influence racial disparities in cancer patient outcome. Additionally, as cancers that have undergone WGD events exhibit unique genetic vulnerabilities, therapies that selectively target WGD-positive cancers may be particularly effective at treating aggressive malignancies in Black patients.

## INTRODUCTION

The unequal landscape of healthcare outcomes in the United States, particularly among Black patients, demands immediate attention. Black patients face higher rates of chronic disease, higher incidence of preventable illness, and higher all-cause mortality compared to white patients^1–4^. These disparities are particularly evident within the field of oncology. For many cancer types, Black patients are diagnosed with more advanced or aggressive cancers than white patients^5–7^. Although cancer outcomes have drastically improved over the past several decades, Black patients continue to have higher rates of cancer-related death^8–10^. Mortality disparities are exacerbated within certain cancer types, including breast and endometrial cancers, in which Black patients exhibit 41% and 21% higher mortality, respectively, compared to white patients^11–13^. Uncovering the mechanisms that mediate these unequal outcomes could help identify prevention or treatment strategies that may aid those populations most at risk.

The source of racial disparities in cancer outcomes is at present unresolved and may result from social, environmental, and genetic factors^14–20^. In the United States, Black individuals are more than twice as likely to live below the federal poverty line than white individuals, and lower socioeconomic status has been linked with decreased healthcare access and lower cancer screening rates^21–25^. In addition, people within economically-deprived communities have disproportionately higher rates of environmental carcinogen exposure, which may increase the risk of cancer development^25,26^. Within the healthcare system itself, Black patients received worse care compared to white patients in more than half of quality care metrics in the 2022 National Healthcare Quality and Disparity Report^27^. These findings extend to cancer-specific interventions, as Black patients experience more delays in chemotherapy induction for breast cancer treatment and are less likely to have adequate oncological resection for gastrointestinal cancers treated with curative intent surgery^28,29^.

In addition to these social and environmental influences on cancer mortality, recent research has raised the possibility that genetic differences between patient populations could affect the development and/or progression of cancer. To date, most population-based cancer profiling efforts have sought to investigate variability in the prevalence of somatic point mutations between racial and ethnic groups^30–34^. These studies have uncovered certain differences in the frequency of mutations in common oncogenes and tumor suppressors that could impact disease pathogenesis. For example, Black patients have been found to exhibit higher rates of mutations in the tumor suppressor *TP53* compared to white patients, and *TP53* inactivation has consistently been linked with poor prognosis^35–41^. Similarly, Black patients with lung cancer have fewer mutations in the druggable oncogene *EGFR*, which may affect treatment options^42^. Uncovering population-based differences in mutation profiles can aid in clinical assessment and may shed light on strategies to ameliorate outcome disparities.

Comparatively less is known about population-based associations for other types of somatic alterations in cancer beyond single-nucleotide point (SNP) mutations. Notably, chromosomal copy number alterations (CNAs) are pervasive across tumor types and have been linked with disease progression, drug resistance, and poor patient outcomes^43–46^. One recent study examined the prevalence of arm-scale CNAs across a large cohort of cancer patients and found few significant differences between Black and white populations^47^. Population-based differences in many other classes of CNAs have not been previously investigated.

One common type of chromosomal alteration is a whole-genome duplication (WGD) event, in which a cell’s chromosome complement doubles. The causes and consequences of WGDs are poorly understood. Mutations in *TP53* have been consistently associated with WGDs in patient sequencing, and cell culture experiments have demonstrated that loss of TP53 facilitates the outgrowth of cells that have undergone a failed mitosis^48,49^.

Environmental carcinogens like combustion products have been shown to cause point mutations, but a link between these pollutants and WGDs has not been previously demonstrated^50,51^. Other genetic and environmental drivers of WGD events remain obscure^44,52,53^. WGDs enhance tumor adaptability and increase metastatic dissemination, potentially by enhancing tumor heterogeneity and allowing cancers to sample a wider range of karyotypes^52,54–56^. Approximately 30% of tumors exhibit WGDs, but whether patient race or ethnicity is associated with these events is unknown^52^.

## RESULTS

### Tumors from self-reported Black patients display an increased frequency of WGDs

WGD events in cancer are associated with genomic instability and aggressive disease (Fig. 1A)^52,57^. We investigated the frequency of WGD events in cancers from different patient cohorts: MSK-MET (n=13,071 patients), The Cancer Genome Atlas (TCGA) (n=8,060 patients), and the Pan-cancer Analysis of Whole Genomes (PCAWG) (n=1,963 patients)^58–60^. These three datasets represent the largest publicly-available tumor sequencing cohorts with both copy number alteration data and patient demographic information. Full demographics of included patients and cancer types can be found in Table S1. We compared the frequency of WGD events in cancers from self-reported Black and white patients in the MSK-MET and TCGA cohorts and between cancers from individuals with inferred African and European ancestry in the PCAWG cohort. We discovered that cancers from self-reported Black patients and patients with inferred African ancestry exhibited a significantly higher incidence of WGDs compared to cancers from self-reported white patients or patients with inferred European genetic ancestry (Fig. 1B-D). Notably, CNAs in each of these datasets were detected using different genomic technologies: SNP arrays, targeted gene sequencing, and whole-genome sequencing, for TCGA, MSK-MET, and PCAWG, respectively^58–60^. As these findings were consistent across all three cohorts, we anticipate that they are robust and independent of any platform-specific artifacts.

**Figure 1.**
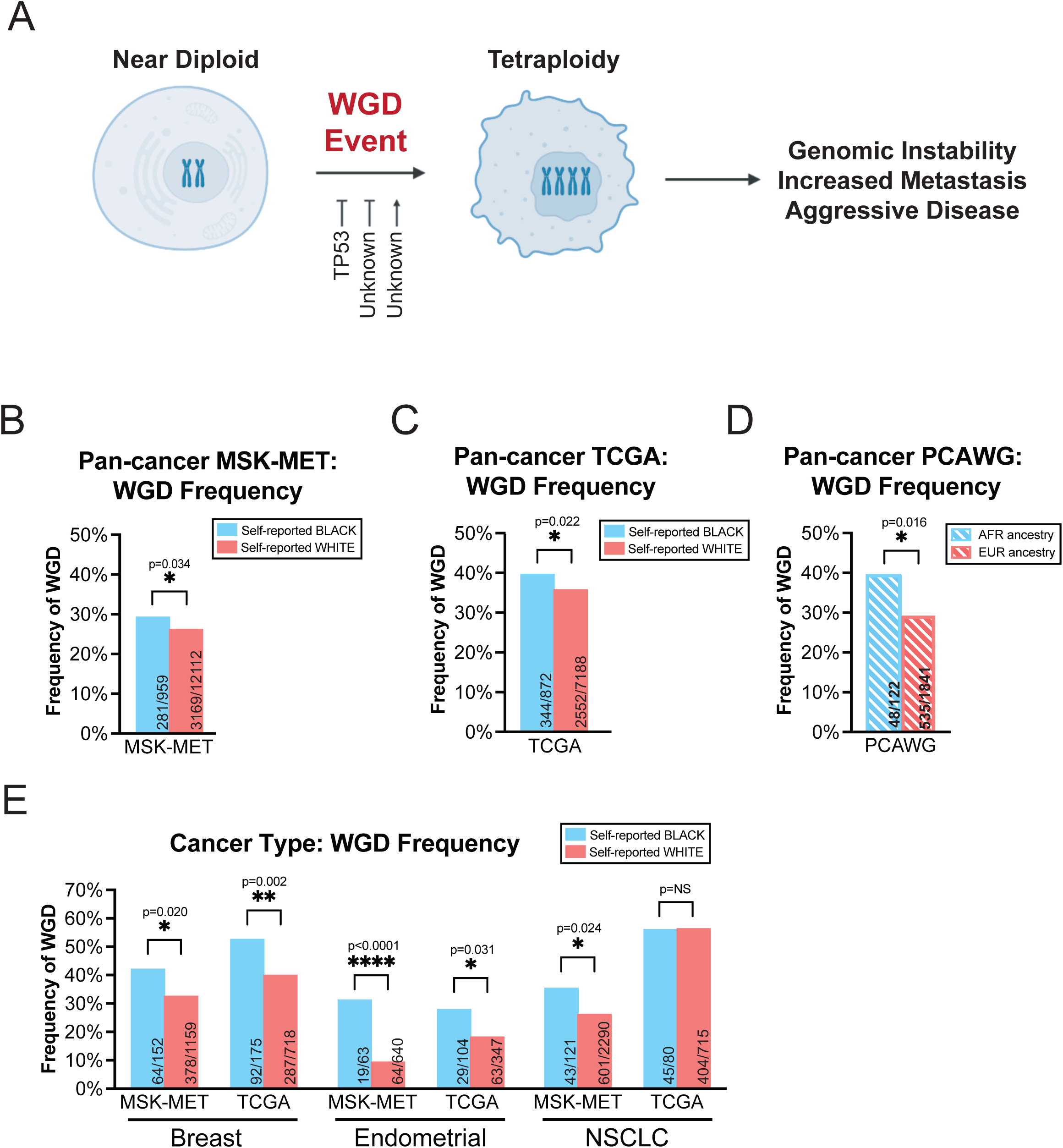
WGDs are more common among self-reported Black cancer patients and those with inferred African genetic ancestry. **(A)** A schematic of the whole-genome duplication process. WGDs produce a cell with a doubled chromosome complement (typically 4N). WGD events are associated with metastasis and disease progression. Loss of the tumor suppressor *TP53* promotes WGDs; other causes of WGDs are largely unknown. **(B-D)** The frequency of WGDs in self-reported Black and white patients from three different cohorts: **(B)** MSK- MET, **(C)** TCGA, and **(D)** PCAWG. Note that in the PCAWG dataset, a patient’s self-reported race was not available, and instead inferred genetic ancestry was used (African (AFR) vs European (EUR)). **(E)** The frequency of WGD events in self-reported Black and white patients with either breast cancer, endometrial cancer, or NSCLC, in the MSK-MET and TCGA cohorts. Statistical testing was performed via two-tailed Pearson’s Chi-squared test. Statistical significance: NS p ≥ 0.05, * p < 0.05, ** p < 0.01, *** p < 0.001, **** p < 0.0001.

The increase in WGD events in self-reported Black patients compared to self-reported white patients ranged from 11% in the TCGA cohort to 35% in the PCAWG cohort. Additionally, the overall rate of WGD events ranged from 26% in MSK-MET to 36% in TCGA. However, within cancer types that were shared across datasets, the frequency of WGD events was highly correlated (R^2^=0.62-0.91, p<0.001-0.0001) (Fig. S1). For instance, thyroid cancers consistently displayed the lowest incidence of WGDs (0-4%) while ovarian cancers consistently displayed the highest incidence of WGDs (55-60%), in alignment with other reports^44,52^. The overall differences in WGD frequency between cohorts may reflect differences in the distribution of cancer types.

Next, we sought to evaluate whether other minority populations also exhibited an increase in WGD events. We therefore investigated whether WGD events were elevated in cancers from patients who identified as Asian, which represented the next-most common racial group in the TCGA and MSK-MET cohorts. In both patient cohorts, we did not detect a significant difference in WGD frequency between self-reported white and Asian patients (Fig. S2A and Table S2). Additionally, tumors from males and females displayed equivalent frequencies of WGD events within each cohort (Fig. S2B, Table S2).

### Tumors from self-reported Black patients with specific cancer types display an increased frequency of WGDs

We considered the possibility that the increased incidence of WGD events in our pan-cancer analysis of self-reported Black patients could result from differences in the representation of distinct cancer types between populations. However, Black patients have historically been underrepresented in genomic studies, which limits our statistical power to detect significant differences in every cancer lineage^61,62^. MSK-MET includes 7.3% self-reported Black patients (n=959 patients), TCGA includes 10.8% self-reported Black patients (n=872 patients), and PCAWG includes 6.2% patients with inferred African ancestry (n=122 patients) (Table S1). To minimize false negatives resulting from the underrepresentation of Black patients, we focused our analysis on cancer types for which we had genomic data from at least 60 self-reported Black patients in both the MSK-MET and TCGA datasets. The three cancer types exceeding this threshold were breast cancer, endometrial cancer, and non-small cell lung cancer (NSCLC) (Table S1). Stratification of PCAWG by cancer type was not performed due to the low number of patients with inferred African ancestry. In both TCGA and MSK-MET, we detected a significant increase in WGD events in self-reported Black patients with breast and endometrial cancers (Fig. 1E). For NSCLC, we detected a significant increase in the MSK-MET cohort but not the TCGA cohort. The increase ranged from 33% in breast cancer (TCGA) to 202% in endometrial cancer (MSK-MET) (Fig. 1E). These results indicate that self-reported Black patients have a significantly higher incidence of WGD events both across cancers and within individual cancer types.

### Analysis of WGD frequency by tumor stage and histological subtype

WGD abundance varies between histological cancer subtypes and is more common in advanced malignancies^52^. Accordingly, we considered the possibility that differences in the prevalence of histological subtypes or differences in the stage at which tumors were diagnosed could produce the increase in WGD events in self-reported Black cancer patients that we observed. However, we determined that WGD events were still significantly more common in tumors from self-reported Black patients for many individual cancer stages and subtypes (Fig. S3-4; Table S1). For instance, self-reported Black patients with either stage II or III breast tumors had a higher incidence of WGD events compared to white patients with similarly-staged tumors (Fig. S3B). Additionally, while the frequency of histological subtypes varied between self-reported Black and white patients, when we limited our analysis to only the most common histological subtypes of each cancer (breast cancer: invasive ductal carcinoma; endometrial cancer: endometrioid tumors, and NSCLC: adenocarcinoma), we still detected an increased frequency of WGD events among self-reported Black patients (Fig. S4A-B). These results indicate that the increased incidence of WGDs in self-reported Black cancer patients is not simply due to differences in tumor stage or histological subtype at diagnosis.

### Eliminating overlap between the TCGA and PCAWG cohorts

596 patients in TCGA were also profiled as part of the PCAWG cohort. To ensure statistical independence, we repeated our analysis of WGD frequencies in TCGA after eliminating the patients who were also included in PCAWG. Within this smaller patient population, we still observed a trend towards increased WGDs in self-reported Black patients in a pan-cancer analysis (p<0.07) and a significant increase in WGDs in self-reported Black patients with breast cancer (p<0.005) (Fig. S5 and Table S3).

### Analysis of WGDs by inferred genetic ancestry

Recent studies have urged caution in the use of race when conducting population-based research, noting that widely used definitions of race represent artificial social constructs^63,64^. Instead, shared genetic ancestry may better reflect population differences in disease risk. Nonetheless, race may still be useful for investigating patterns of heath and disease in the US due to its association with the social determinants of health, including poverty, pollution, systemic racism, and lack of healthcare access^65^. According to the National Academy of Sciences’ 2023 report *Using Population Descriptors in Genetics and Genomics Research*, “race… may be a useful population descriptor for researchers who wish to measure a consequential form of social status and affiliation… [R]ace may be a proxy for the experience of racism in health disparities studies.”

As described above, we found that both individuals who self-reported as Black or African American and individuals with African ancestry were more likely to exhibit WGD-positive cancers compared to individuals who self-reported as white or individuals with European ancestry (Fig. 1B-E). To extend this investigation, we re-analyzed the MSK-MET dataset using inferred genetic ancestry instead of racial self-reporting^34^. We found that the overall patterns of WGD were conserved: African ancestry was associated with a 16% increase in WGD events in a pan-cancer analysis and with a 32-135% increase in breast, endometrial, and NSCLC (Fig. S6A-B; Table S4). Additionally, inferred African ancestry and inferred European ancestry were 96.3% and 99.9% concordant with self-reported Black and white identity, respectively (Fig. S6C-D). Finally, the fraction of inferred African ancestry was associated with an increasing frequency of WGDs, while the fraction of inferred European ancestry was associated with a decreasing frequency of WGDs (Fig. S6E-F; p=0.050 and p=0.002, respectively). We conclude that WGD events are associated with both inferred African ancestry and Black self-identity.

### Increasing aneuploidy burden associated with WGD events in self-reported Black patients

Previous analyses of genetic differences between cancers from Black and white patients have focused on exploring the spectrum of somatic point mutations in each population^30–34,42^. As our investigation uncovered a significant increase in the frequency of WGDs in Black patients, we next sought to expand our analysis of CNAs to interrogate all aneuploidy events. We calculated the aneuploidy burden, defined as the sum total of arm-scale CNA events, in each tumor genome. We discovered that cancers from self-reported Black patients had higher levels of aneuploidy in a pan-cancer analysis and in the breast and endometrial subtypes (Fig. S7A-C). However, cancers that have undergone WGD events have consistently been observed to exhibit a higher aneuploidy burden than WGD-negative cancers^56,66^. Indeed, when we analyzed the aneuploidy levels in WGD-positive and WGD-negative cancers separately, the differences between Black and white patients were muted (Fig. S7D-F). There was no significant increase in aneuploidy burden among WGD-negative Black patients, and among WGD-positive Black patients there was an increase in aneuploidy burden in the TCGA cohort but not the MSK-MET cohort in a pan-cancer analysis.

Finally, we examined individual chromosome arm gain and loss events in self-reported Black and white cancer patients. We found that cancers from self-reported Black patients were significantly more likely to lose the q arm of chromosome 4 and p arm of chromosome 8 compared to cancers from self-reported white patients (Fig. S7G). There was no significant difference among any of the other 39 possible aneuploidies that were quantifiable. In a subtype-specific analysis, we did not observe any consistent difference in aneuploidy patterns in NSCLC, while in breast and endometrial cancers, self-reported Black patients demonstrated increased frequencies of specific aneuploidy events shared between MSK-MET and TCGA. In breast cancer, self-reported Black patients demonstrated an increase in 16q gains compared to self-reported white patients. Similarly, self-reported Black patients consistently displayed 16q and 17p loss events in endometrial cancer. When patients were subdivided by WGD status, no significant differences in arm-scale aneuploidies were detected (Fig. S7G). We conclude that there are moderate differences in the quantity and frequency of arm-scale aneuploidies in self-reported Black patients, though the most prevalent and consistent copy number difference is an increase in WGD events.

### Association of WGD events with TP53 mutations

We sought to uncover the somatic alterations associated with WGD events in self-reported Black and white patients. For this and subsequent analyses, we focused on the MSK-MET cohort, as this was the largest single patient cohort and had the best sequencing coverage of cancer-relevant genes. We constructed a logistic regression model linking somatic alterations with WGD status while correcting for cancer type (Fig. 2A, Table S5). The strongest predictor of WGD status was the presence of mutations in *TP53*^52^. In contrast, mutations in *KRAS*, *BRAF*, and *PTEN* were significantly associated with a reduced likelihood of WGD events. The association between *TP53* mutations and WGD events has been previously observed, as loss of p53 enhances the proliferation of cells that have undergone tetraploidization^52,67,68^. The reasons why mutations in certain other oncogenes and tumor suppressors are associated with fewer WGD events is at present unclear and may reflect differences in the evolutionary trajectories of these cancers^69,70^. We repeated our logistic regression analysis for self-reported Black and white patients, considered separately (Fig. 2B-C, Table S6-7). In self-reported Black patients, the only significant features associated with WGD status with a q-value <0.1 were *TP53* mutations and amplifications of cyclin E (*CCNE1*). Cyclin E gains have recently been identified as a driver of WGDs, and both features were also significantly associated with WGD events in our logistic regression model of tumors from white patients (Fig. 2C)^53^. These results suggest that similar somatic alterations drive WGDs in both self-reported Black and white patients.

**Figure 2.**
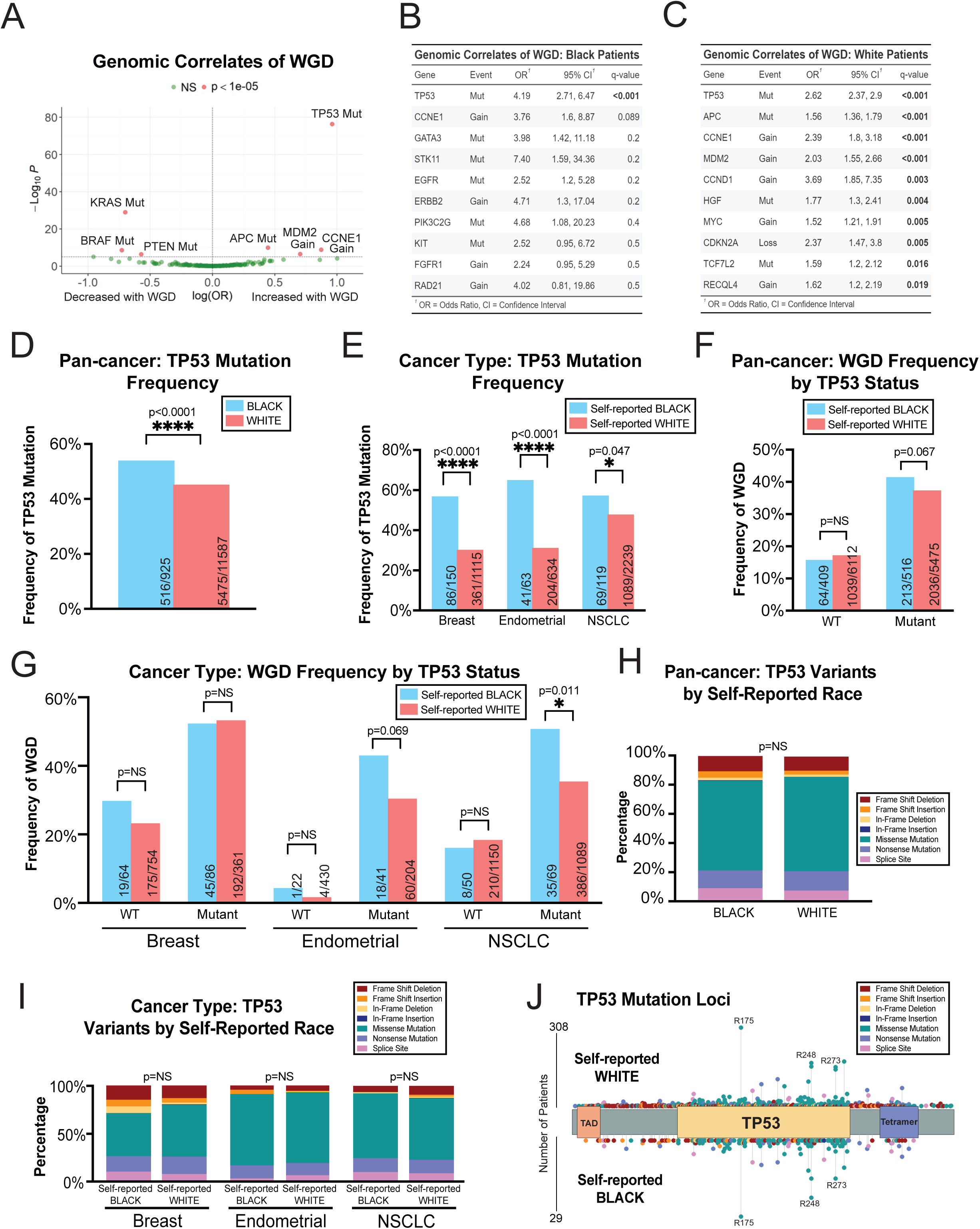
Genetic analysis of WGD events in self-reported Black and white cancer patients. **(A)** A volcano plot displaying genetic alterations associated with an increased or decreased likelihood of WGD events across both self-reported Black and white patients in the MSK-MET cohort. Acronyms: Mutant (Mut). **(B)** A table displaying 10 events exhibiting the strongest correlation with WGD events among self-reported Black patients. **(C)** A table displaying 10 events exhibiting the strongest correlation with WGD events among self-reported white patients. **(D)** A bar graph displaying the frequency of *TP53* mutations across self-reported Black and white cancer patients. **(E)** A bar graph displaying the frequency of *TP53* mutations across self-reported Black and white patients with either breast cancer, endometrial cancer, or NSCLC. **(F)** A bar graph displaying the frequency of WGD events among self-reported Black and white cancer patients, divided based on *TP53* status. **(G)** A bar graph displaying the frequency of WGD events among self-reported Black and white patients with either breast cancer, endometrial cancer, or NSCLC, divided based on *TP53* status. **(H)** A bar graph displaying the distribution of different types of *TP53* mutations in self-reported Black and white cancer patients. **(I)** A bar graph displaying the distribution of different types of *TP53* mutations in self-reported Black and white patients with either breast cancer, endometrial cancer, or NSCLC. **(J)** A lollipop plot displaying the sites of TP53 mutations in tumors from either self-reported white (top) or Black (bottom) cancer patients. Statistical testing was performed via Wald test (A-C) with correction by Benjamini-Hochberg’s method (B-C) and two-tailed Pearson’s Chi-squared test (D-I). Statistical significance: NS p ≥ 0.05, * p < 0.05, ** p < 0.01, *** p < 0.001, **** p < 0.0001.

As the types of somatic alterations associated with WGD events in self-reported Black and white patients were similar, we considered the possibility that differences in frequencies of these alterations could influence the prevalence of WGDs. Consistent with previous observations, we found that self-reported Black patients were significantly more likely to harbor *TP53* mutations than self-reported white patients in both a pan-cancer analysis and in breast and endometrial cancers but not NSCLC (Fig. 2D-E)^35–40^. Next, we separated patients based on *TP53* status. We observed that there was no significant difference in WGD frequency between Black and white patients with *TP53*-WT cancers. However, among patients with *TP53*-mutant cancers, self-reported Black patients showed a trend towards increased WGD events in a pan-cancer analysis (p=0.067) and a significant increase in the NSCLC cohort (p=0.011) (Fig. 2F-G). These results suggest that the increased frequency of *TP53* mutations contributes to but cannot fully account for the increased incidence of WGD events in Black patients.

Next, we investigated whether different classes of *TP53* mutations could underlie the different prevalence of WGD events within self-reported Black and white populations. However, the overall distribution of classes of *TP53* mutations (e.g., missense vs nonsense) was similar between self-reported Black and white patients (Fig 2H-I). Additionally, the same set of p53-inactivating point mutations, including the R175, R248, and R273 mutations, were commonly observed among both Black and white patients (Fig. 2J).

We subsequently investigated the association between WGD events and CCNE1 amplifications in self-reported Black and white patients. Consistent with previous results, we found that CCNE1 amplifications were more common in self-reported Black patients (Fig. S8A-B)^35^. However, when we split patients based on CCNE1 amplification status, we observed that a consistent difference in the frequency of WGD events between self-reported Black and white patients remained (Fig. S8C-D). We conclude that somatic genetic alterations are unable to fully account for increased prevalence of WGD events among self-reported Black patients.

### Association between self-reported race, WGD Status, and patient outcome

We sought to determine whether the increased incidence of WGD events in self-reported Black patients was linked with racial disparities in patient outcome. Consistent with previous observations, WGD events were strongly associated with hallmarks of aggressive disease^52,71^. The rate of WGDs was significantly higher in metastatic samples compared to primary tumor samples in both a pan-cancer analysis and in individual cancer types (Fig S9A-B). WGD-positive tumors were more likely to exhibit progressive disease after frontline treatment and patients with WGD-positive tumors were less likely to be tumor-free at the conclusion of the observational follow-up period (Fig S9C-D). We performed a multivariate Cox proportional hazard regression analysis including WGD status and common clinical variables (age, sex, *TP53* status, MSI status, aneuploidy burden, cancer type). WGD status was significantly associate with shorter patient survival even including these known disease-modifying factors [HR 1.21 (1.12-1.30), p<0.001] (Fig S10). These results are in agreement with other studies demonstrating that WGD can drive malignant progression^52,72^.

Next, we looked at the interaction of self-reported race, WGD, and patient outcome. Consistent with known national trends, we found that self-reported Black cancer patients exhibited a significantly shorter overall survival time following diagnosis compared to self-reported white patients (Fig. 3A)^8–10^. Similarly, WGD events were also associated with worse patient outcomes in a Kaplan-Meier analysis (Fig. 3B). Interestingly, among the subset of patients with WGD-positive tumors, there was no significant difference in survival time between self-reported Black and white patients (Fig. 3C). However, within the WGD-negative patient subset, a significant difference in survival between self-reported Black and white patients was observed (Fig. 3D). These results are consistent with prior reports demonstrating that WGD events drive metastases and aggressive disease, and suggest that the increased incidence of WGD events in self-reported Black patients may be linked with worse overall survival^46,58,73^. We speculate that within WGD-negative patients, WGD events may be occurring post-diagnosis, or additional genetic and environmental factors may be contributing to these disparate outcomes.

**Figure 3.**
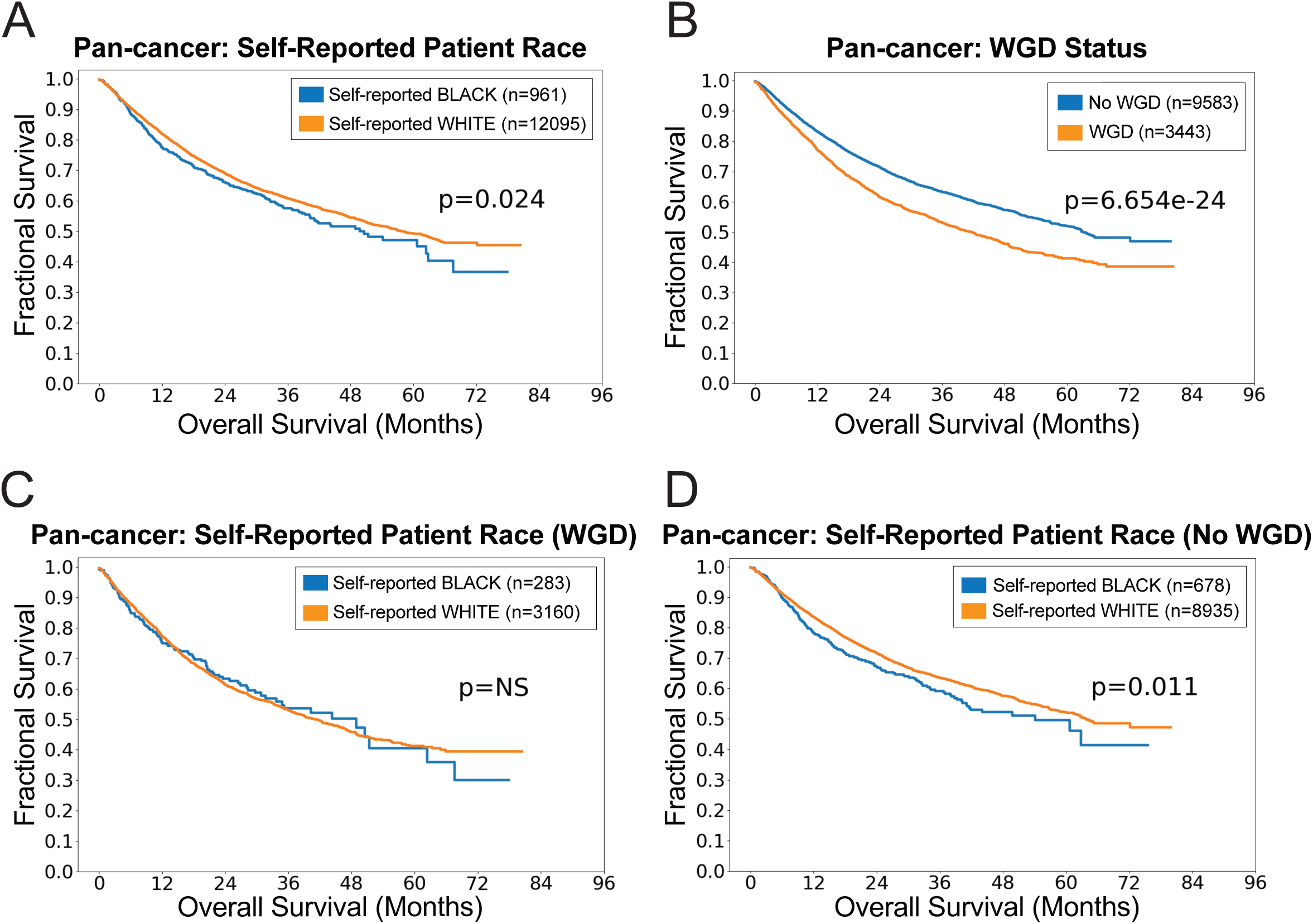
Pan-cancer survival analysis in self-reported Black and white cancer patients. **(A)** A Kaplan-Meier plot displaying survival of self-reported Black and white patients in the MSK-MET cohort. **(B)** A Kaplan-Meier plot displaying survival of cancer patients based on WGD status. **(C)** A Kaplan-Meier plot displaying survival of WGD-positive self-reported Black and white cancer patients. **(D)** A Kaplan-Meier plot displaying survival of WGD-negative self-reported Black and white cancer patients. Statistical testing was performed via logrank test (A-D).

We repeated this analysis within the MSK-MET breast, endometrial, and NSCLC cohorts. We observed that self-reported Black patients with endometrial cancer, but not breast or NSCLC, had significantly shorter survival times compared to white patients within this study (Fig. S11A-C). Racial disparities within breast and NSCLC outcomes have been reported in other analyses, and we speculate that the MSK-MET cohort sizes may not be sufficiently large to detect survival differences that are moderate overall^11–13^. Within endometrial cancer, we observed that patients with WGD-positive tumors had worse outcomes (Fig. S11D). Consistent with our pan-cancer analysis, within the subset of tumors that were WGD-positive there was no significant difference between self-reported Black and white patient outcomes, while a significant difference remained apparent within the WGD-negative subset (Fig S4E-F).

Finally, we further divided the pan-cancer survival analysis based on *TP53* status. As expected, *TP53*-mutant tumors had significantly worse overall survival (Fig. S12A). In general, further subdividing patients based on *TP53* status, in addition to race and WGD status, minimized race-based differences in survival time (Fig. S12B-I). In total, these results suggest that the increased incidence of *TP53* mutations and WGD events contribute to the worse overall outcomes for self-reported Black cancer patients, although our findings do not rule out the influence of additional social, environmental, and genetic factors.

### Analysis of WGD events in self-reported Black and white patients with prostate cancer

In the United States, there is a pronounced and pernicious racial disparity between Black and white patients with prostate cancer^74,75^. Overall, African Americans are about twice as likely to have metastatic disease and die of prostate cancer compared to white Americans^76,77^. We therefore investigated whether WGD events could contribute to this disparity. We found that there was no significant difference in WGD frequency between self-reported Black and white patients in the MSK-MET cohort (Fig. S13A). We obtained a similar result within TCGA, though we note that this analysis is limited by the fact that there are only seven self-reported Black patients with prostate cancer in this cohort (Fig. S13B). Consistent with previous studies, we found that self-reported Black prostate cancer patients had worse overall survival than white patients and WGD events were also associated with shorter survival (Fig. S13C-D)^74,77^. As with our pan-cancer analysis, we found that the survival disparities between Black and white patients was maintained among WGD-negative cancers, while no difference in survival was apparent among cancers that have undergone WGDs (Fig. S13E-F). In total, our data indicate that WGD events are associated with aggressive disease in prostate cancer overall; however, these data also suggest that WGD events themselves are not a prominent driver of disparities in this specific cancer type.

### Self-reported race and WGD status are associated with metastases to the same anatomic sites

Next, we used additional patient information that was collected as part of the MSK-MET dataset to further explore the clinical correlates of self-reported race and WGD status. Within this cohort, self-reported Black patients were diagnosed and underwent surgery at earlier ages compared to self-reported white patients, and self-reported Black patients were also more likely to die younger (Fig. 4A). Similarly, WGD-positive tumors were associated with younger diagnoses, younger age at surgery, and younger death in all patients, regardless of race (Fig. 4B). Next, we examined microsatellite instability in each patient, which is a genomic state characterized by the accumulation of point mutations in repetitive sequences^78,79^. We found that tumors from self-reported Black patients were significantly less likely to exhibit high microsatellite instability (MSI-H) compared to tumors from white patients, and MSI-H status was also less common in WGD-positive tumors (Fig. 4C-D). MSI-H status has been linked with favorable outcomes, and the under-representation of MSI-H cancers among tumors from both self-reported Black patients and patients with WGD-positive disease could represent another factor that contributes to differences in patient survival^80,81^.

**Figure 4.**
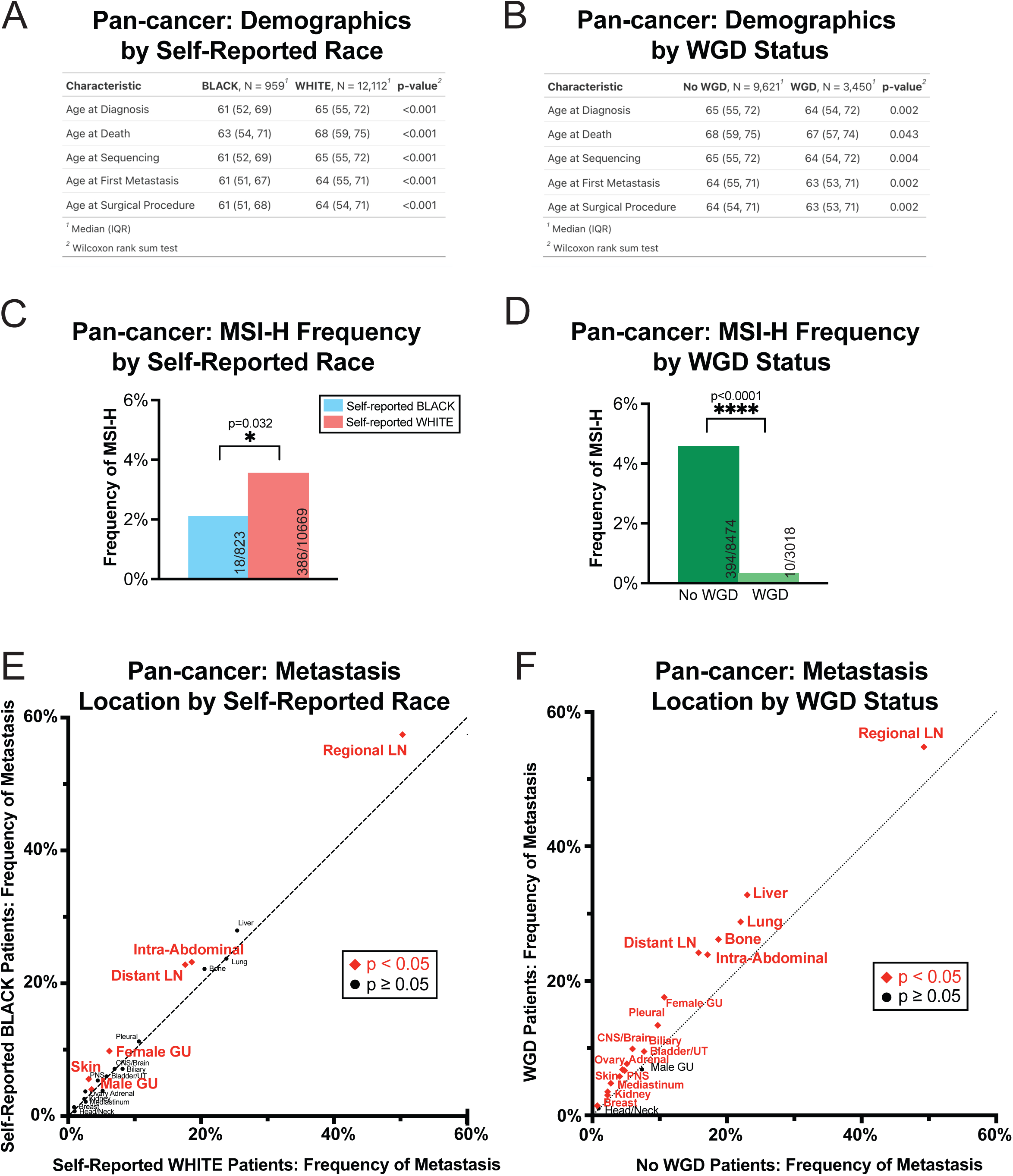
Clinical correlates of WGD status in self-reported Black and white cancer patients. **(A)** A table displaying the demographics of self-reported Black and white cancer patients in the MSK-MET cohort. **(B)** A table displaying the demographics of cancer patients based on WGD status. **(C)** A bar graph displaying the frequency of microsatellite instability (MSI-H) among tumors from self-reported Black and white cancer patients. (**D)** A bar graph displaying the frequency of microsatellite instability (MSI-H) among cancer patients based on WGD status. **(E)** The frequency of metastatic dissemination to different anatomic sites, divided by patient race. Locations written in red are significantly more likely among self-reported Black patients, no locations were more likely among self-reported white patients. For Ovary and Female Genital locations, frequency of metastasis represents female patients only. For Male Genital location, frequency of metastasis represents male patients only. **(F)** The frequency of metastatic dissemination to different anatomic sites based on WGD status. Locations written in red are significantly more likely among WGD-positive patients, no locations were more likely among WGD-negative patients. For Ovary and Female Genital locations, frequency of metastasis represents female patients only. For Male Genital location, frequency of metastasis represents male patients only. Acronyms: Central Nervous System (CNS), Lymph Node (LN), Peripheral Nervous System (PNS), Urinary Tract (UT). Statistical testing was performed via Wilcoxon rank-sum test (A-B) and two-tailed Pearson’s Chi-squared test (C-F). Statistical significance: NS p ≥ 0.05, * p < 0.05, ** p < 0.01, *** p < 0.001, **** p < 0.0001.

Finally, we compared the frequency of metastatic dissemination to different anatomic sites between patient populations. Consistent with established disparities in overall outcomes, we observed that self-reported Black cancer patients had a greater incidence of metastatic disease compared to white patients. Notably, we found that self-reported Black patients had significantly higher rates of metastases to regional lymph nodes, distant lymph nodes, intra-abdominal space, the male and female genitourinary system, and skin (Fig. 4E, Table S8-9). Intriguingly, metastases to each of these sites except the male genitourinary system was also more common among WGD-positive tumors compared to WGD-negative tumors (Fig. 4F, Table S10-11). In total, these findings suggest that several differences in the clinical presentation of cancers in self-reported Black and white populations could be related to the increased incidence of WGDs among Black patients.

### Carcinogen exposure drives WGD events in cell culture

As we found that the genetic drivers of WGD events were highly similar between self-reported Black and white patients, we next sought to investigate whether differences in environmental exposures could contribute to the increased WGD events observed in self-reported Black patients. In the United States, Black Americans are disproportionately exposed to environmental carcinogens^82–84^. This has been partially attributed to historic redlining, a discriminatory practice in which minority communities were concentrated in less desirable neighborhoods near pollution-emitting factories and highways^85,86^. In contemporary epidemiological studies, minority communities continue to live in disadvantaged neighborhoods with higher rates of pollution exposure and cancer mortality compared to white Americans^87,88^. Many carcinogens are known to accelerate the development of point mutations; however, a link between environmental pollutants and WGD events has not been reported^50,51^.

We established a cell culture system to investigate the link between various common carcinogens and WGDs^89,90^. We cultured murine lung epithelial cells alone (monoculture system) or, as a model of lung inflammation, we cultured murine lung epithelial cells along with alveolar macrophages (co-culture system) (Fig. 5A). We exposed the monoculture and co-culture systems to a selection of common carcinogens, including combustion products, carbons, clays, and various metal oxides, and performed live-cell imaging to follow mitotic progression (Fig. 5B, Table S12). We applied concentrations of carcinogens that were not overtly toxic, as the overall rates of mitotic division were largely unaffected by carcinogen exposure (Fig. 5C, Fig. S14A). In the monoculture system, 0 out of 13 tested carcinogens caused an increase in binucleation events (Fig. S14B). However, in the co-culture system, 5 out of 13 carcinogens, including 4 out of 4 combustion products, caused a significant increase in binucleation events (Fig. 5D). For instance, exposure to Printex90, a model for carbon particles found in soot as a result of the combustion of coal tar, petroleum, or other carbon-based materials, caused a 9% increase in binucleation events during a 24 hour period (p=0.003)^91,92^. In total, these results suggest that common environmental carcinogens not only drive the development of point mutations but can also promote WGDs by triggering mitotic failure and binucleation.

**Figure 5.**
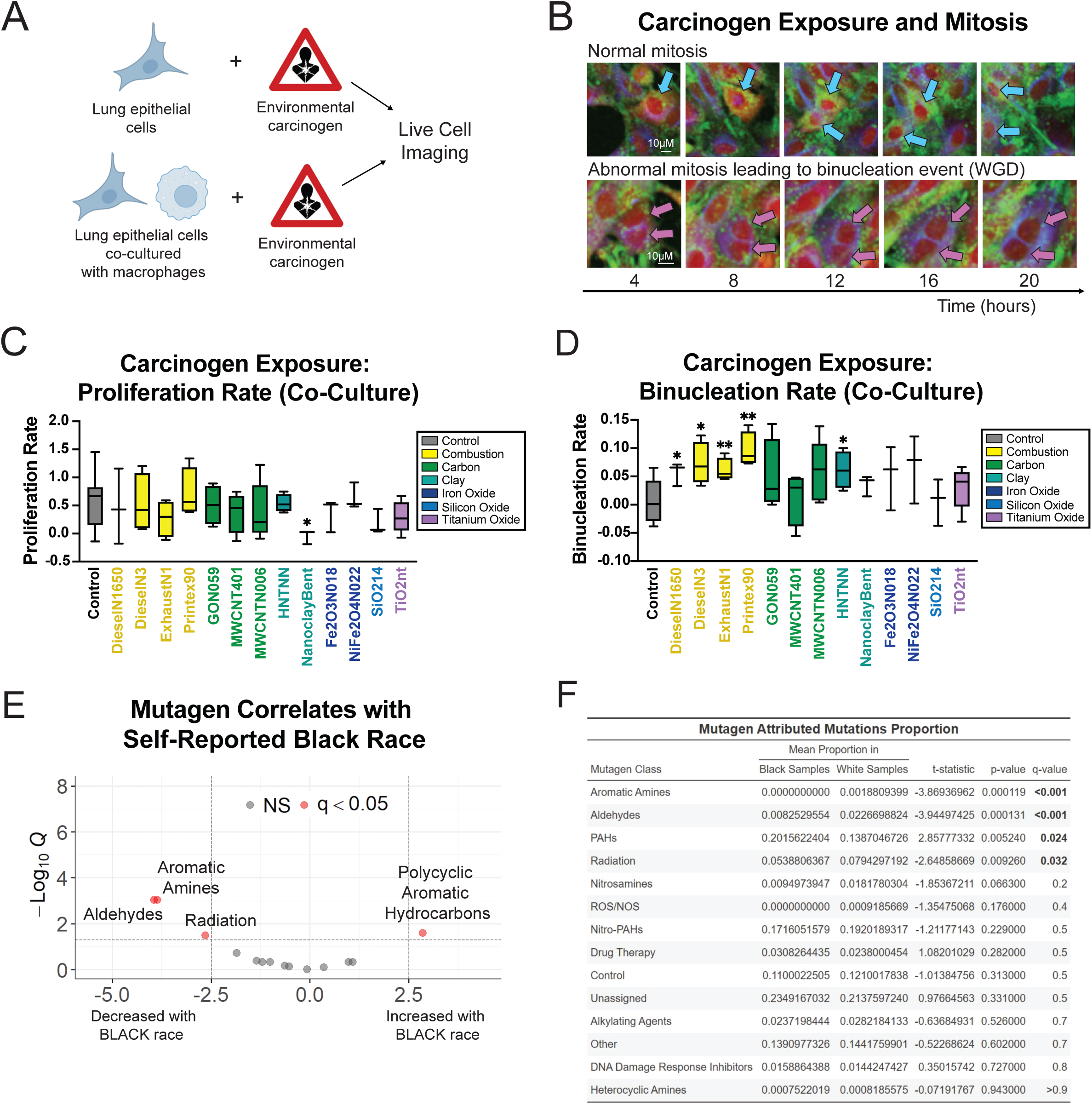
Carcinogen exposure effects on mitosis. **(A)** A schematic outlining the experimental design. **(B)** Live cell microscopy images to illustrate examples of normal mitosis and abnormal mitosis leading to binucleation events (WGD) for quantification after carcinogen exposure. **(C)** A box plot displaying the proliferation rate of lung epithelial cells co-cultured with alveolar macrophages after exposure to various carcinogens. **(D)** A box plot displaying the proliferation rate of lung epithelial cells co-cultured with alveolar macrophages after exposure to control and various carcinogens. Statistical testing was performed via pairwise two-tailed t-tests to reference control (C-D). **(E)** A volcano plot showing the mutagen class attributed proportions associated with self-reported racial group. Difference in the proportion of mutagen-attributed mutations between self-reported Black and white racial groups were tested by two-tailed Student’s t-tests followed by Benjamini-Hochberg correction^135^. Red points indicate q-value < 0.05. **(F)** A table displaying proportions mutagen-attributed mutations by mutagen signature classes between self-reported Black and white patients. Difference in proportion of mutagen-attributed mutations between self-reported Black and white racial groups were tested by two-tailed Welcht’s t-tests followed by Benjamini-Hochberg correction^135^. Acronyms: polycyclic aromatic hydrocarbons (PAHs), reactive oxygen species (ROS), nitric oxide species (NOS). Statistical significance: NS p ≥ 0.05, * p < 0.05, ** p < 0.01, *** p < 0.001, **** p < 0.0001.

### Signatures of carcinogen exposure in lung tumors from self-reported Black and white patients

We speculated that differences in environmental carcinogen exposure could influence the development of WGDs in Black and white cancer patients. While nationwide disparities in pollution exposure between Black and white Americans are well-documented, we do not know the exposure histories of the patients in our sequencing cohorts^93–95^. In order to investigate whether Black and white patients in our cohorts exhibited evidence of differential pollution exposure, we analyzed carcinogen-associated mutational signatures in the NSCLC tumors from TCGA. Based on prior work, we established signatures for classes of common mutagens including radiation, alkylating agents, and heterocyclic amines^96^. Interestingly, we found that lung tumors from self-reported Black patients exhibited a significant increase in the proportion of mutations associated with polycyclic aromatic hydrocarbon (PAH) exposure (Fig. 5E-F). PAHs are a common urban pollutant produced by burning carbon, and combustion products like Printex90 and diesel exhaust are significant sources of PAH exposure^97–100^. In contrast, lung tumors from self-reported white patients exhibited evidence of higher radiation and aldehyde exposure (Fig. 5E-F). While we do not know the specific exposure histories of these patients, this analysis suggests that Black patients may be exposed to different environmental pollutants than white patients. Notably, we found evidence of a combustion-associated mutational signature in lung tumors from self-reported Black patients, and we found that combustion byproducts were sufficient to trigger mitotic failure in cultured lung epithelial cells.

## DISCUSSION

In this work, we found that self-reported Black or African American cancer patients exhibited a significantly greater incidence of WGD events compared to white cancer patients. This discrepancy was detectable in both a pan-cancer analysis and in several individual cancer subtypes. Historically, most research on genetic differences between cancer patient populations has focused on single-nucleotide point mutations; our work demonstrates the existence of significant, outcome-associated differences in patterns of chromosomal alterations as well^30–33^. We speculate that analyzing other types of genetic or epigenetic alterations in cancer (e.g., methylation patterns, smaller CNAs, intratumoral heterogeneity, etc.) may reveal additional informative differences^101^.

Our work is consistent with previous reports that documented an increased incidence of WGD events among African ancestry prostate cancer patients in sub-Saharan Africa^102,103^. However, our analysis lacked sufficient numbers to recapitulate this finding, as the TCGA cohort only included seven Black patients with prostate cancer (Fig. S13, Table S1). More broadly, while African American patients are significantly underrepresented in most genomic studies, dedicated sequencing efforts specifically designed to assess underrepresented patient populations have uncovered a wealth of new cancer drivers and vulnerabilities, illustrating the power of these focused efforts. Notably, the cancer types in which self-reported Black patients exhibit frequent WGD events (breast, endometrial, prostate, and NSCLC) are also among those that have been recognized as exhibiting the most significant racial disparities in patient incidence or outcome^13,19,20,33^. Furthermore, we found that, among WGD-positive cancer patients, there were no differences in overall survival times, suggesting that WGDs may represent one mechanism underlying the disparate outcomes that have been demonstrated within the American healthcare system.

Our genetic analysis of tumor sequencing data revealed a strong association between *TP53* mutations and WGD events in both self-reported Black and white patients. Consistent with previous results, we found that Black patients exhibited a higher incidence of *TP53* mutations overall^35,36,38^. However, WGD events were still more common among self-reported Black patients with *TP53*-mutant cancers compared to self-reported white patients. Our work did not identify any significant differences in the genetic drivers of WGDs associated with patient race, suggesting that these events may be influenced by epigenetic or environmental factors. Furthermore, our preliminary evidence demonstrates that carcinogen exposure, particularly combustion agents, result in WGD events in vitro, thereby providing a potential mechanistic link between known social determinants of health and aggressive disease. This is particularly important because African Americans in the US are more likely to live in areas with higher rates of carcinogenic air pollution, including diesel exhaust, which could affect the development of WGDs in lung cancer^25,26,104^. Additional work will be required to explore the source(s) of the disparate rates of WGDs between patient populations.

The reason why carcinogen exposure triggered mitotic failure only in epithelial cells co-cultured with macrophages is at present unclear. It has previously been observed that epithelial cells are capable of tolerating foreign particulate matter, in part by upregulating lipid metabolism genes and sequestering the particles at the cell membrane. In contrast, the same particulate matter exposure results in cell death in macrophages, which causes the re-release of the particulate matter into the surrounding environment and the secretion of pro-inflammatory cytokines^89^. We speculate that certain paracrine signals from the macrophages may be affecting mitosis in the epithelial cells, though the identity of those signals is at present unknown.

Finally, the increase in WGD events among self-reported Black patients that we have documented has the potential to influence patient staging and treatment. We found that WGD-positive tumors were more likely to have spread to regional or distant lymph nodes, which may warrant additional surveillance and interventions among Black patients. More broadly, chromosomal alterations are strongly associated with patient outcome, and analyzing tumor karyotypes as part of a standard pathological workup may improve our ability to preemptively detect aggressive disease^45,105–109^. Additionally, recent research has demonstrated that WGD-positive cancer cells harbor unique genetic vulnerabilities. For instance, alterations in the mitotic apparatus resulting from WGD events cause cells to become dependent on the mitotic kinesin KIF18A, which is otherwise dispensable in diploid cells^70,110^. AMG650 is a small molecular inhibitor of KIF18A that has entered Phase I clinical trials, underscoring the recent progress toward selectively targeting WGD-positive cancers^111^. Therapies designed to selectively target WGD-positive tumors may be particularly effective in Black cancer patients and could serve to ameliorate the disparate racial outcomes in cancer mortality.

## MATERIALS AND METHODS

### Data Acquisition

Mutational, copy number, sample, and patient data were downloaded from the cBioPortal datahub (https://github.com/cBioPortal/datahub)^112,113^. Whole genome duplication determinations were sourced from the original cohort study (MSK-MET and PCAWG) or from subsequent analyses (TCGA)^44,52,60^. A sample was considered to have undergone WGD based on processing by FACETS (MSK-MET), consensus across 10 different methods (PCAWG), or ABSOLUTE (TCGA)^60,114,115^. Genetic ancestry determinations for the MSK-MET cohort were graciously provided by Kanika Arora, following the method from Arora, et al. (2022)^34^. Racial data was determined by electronic health record review (MSK-MET) and by interview during enrollment where patients were asked to select from the racial categories defined by the U.S. Office of Management and Business and used by the U.S. Census Bureau (TCGA). Regional lymph node metastasis data were sourced from Nguyen, et al. (2022)^58^. Mutagen signatures of environmental agents were from the original study^96,116^.

### Data Harmonization and Cleaning

Data was harmonized across cohorts by including lesions with known WGD status within sample data, considering only genomic aberrations for genes found in the IMPACT-505 geneset within mutational and copy number data, and maintaining patients with known sex, and self-reported race or inferred ancestry within patient data depending on availability of either self-reported race or ancestry data. In MSK-MET, patients with self-reported race of Asian-far east/Indian (Asian), Black or African American (Black), or white and inferred ancestry of either African or European were maintained separately; in TCGA, patients with self-reported race of Asian, Black, or white were maintained; in PCAWG, patients with inferred ancestry of African or European were maintained. With the exception of analyzing WGD frequency in metastatic samples, all included samples were from primary lesions with determinable WGD status, known sex, and had self-reported race of Asian, Black, or white (MSK-MET and TCGA) or inferred genetic ancestry of African or European (MSK-MET and PCAWG). Excluded samples were those from metastatic lesions, without determinable WGD status, unknown sex, or from either a racial group or genetic ancestry other than those explicitly included. Metastatic samples included for supplemental analysis included MSK-MET samples with determinable WGD status, known sex, and had self-reported race of Black or white. To be included in our analysis, genomic aberrations had to occur in ≥200 patients across racial groups and ≥2% of patients within a racial group. We focused mutational analysis on nonsynonymous mutations and trichotomized copy number levels to neutral, loss, and gain. As the cohorts we analyzed do not use the same identifiers for cancer type, we considered all cancer types regardless of identifier in pan-cancer analyses, while standardizing across cohorts for comparisons by cancer type. Standardization was as follows: breast cancer were samples listed as BRCA (TCGA) or Breast Cancer (MSK-MET); endometrial cancer were samples listed as UCEC (TCGA), Endometrioid Adenocarcinoma (MSK-MET), Serous Carcinoma (MSK-MET); non-small cell lung cancer (NSCLC) were samples listed as LUAD (TCGA), LUSC (TCGA), or Non-Small Cell Lung Cancer (MSK-MET); prostate cancer were samples listed as PRAD (TCGA) or Prostate Cancer (MSK-MET). A positive determination for regional lymph node metastasis was made according to the presence of “regional_lymph” in the “met_site_mapped” variable from Nguyen, et al. (2022)^58^. The code used to perform this analysis is available at https://github.com/sheltzer-lab/wgd_disparities.

### Software Versions

Analyses were performed using R (version 4.2.2)^117^, PRISM (version 9.4.1), and Python (version 3.10.8)^118^. R packages: car (version 3.1-2)^119^, EnhancedVolcano (version 1.16.0)^120^, ggplot2 (version 3.4.2)^121^, gt (version 0.9.0)^122^, gtsummary (version 1.7.2)^123^, maftools (version 2.14.0)^124^, openxlsx (version 4.2.5.2)^125^, reshape (version 1.4.4)^126^, tidyverse (version 2.0.0)^127^. Python packages: lifelines (version 0.27.4)^128^, matplotlib (version 3.6.2)^129^, and pandas (version 1.5.2)^130^. Carcinogen analyses and life cell imaging software included Imspector(version 16.2.8282-metadata-win64-BASE) software provided by Abberior Instruments^131^; Fiji, ImageJ 1.52p (NIH)^132^; Mathematica 12.0, license L5063-5112 (Wolfram)^133^. Mutagen signature attributions were determined by signature.tools.lib (v2.4.4)^134^.

### Whole-genome Duplication Frequency Analysis of MSK-MET, TCGA, PCAWG

The frequency of WGD was calculated as the number of WGD-positive samples over the total sample count within a self-reported racial group, inferred ancestry, or self-reported sex at a pan-cancer level and additional subset by cancer type; testing for association was done via two-sided Pearson’s Chi-squared test. WGD frequencies by cancer type were compared between MSK-MET, TCGA, and PCAWG via Pearson correlation.

### Whole-genome Duplication Frequency in TCGA-Exclusive Patients

To analyze independent patients within the TCGA dataset, shared samples between TCGA and PCAWG were removed. The frequency of WGD was calculated as the number of WGD-positive samples over the total sample count (TCGA-Exclusive) within a self-reported racial group for pan-cancer and cancer type analyses; testing for association was done via two-sided Pearson’s Chi-squared test and Fisher’s exact test across racial groups.

### Whole-genome Duplication Frequency Analysis by Metastatic Status and Cancer Stage

The frequency of WGD was calculated as the number of WGD-positive samples over the total sample count within a self-reported racial group by metastatic status (MSK-MET) and stage (TCGA) for each cancer type: testing for association was done via two-sided Pearson’s Chi-squared test. Available staging information in TCGA was utilized which includes pathological staging for breast cancer and NSCLC and clinical staging for endometrial cancer. Metastatic status and cancer staging distributions were summarized within each racial group by cancer type; testing for association was done via two-sided Pearson’s Chi-squared test and Fisher’s exact test across racial groups.

### Whole-genome Duplication Frequency Analysis by Histological Subtype

Shared cancer types between MSK-MET and TCGA were aggregated. For breast cancer, IDC includes MSK-MET histological designations HR+/HER2+ Ductal Carcinoma, HR+/HER2- Ductal Carcinoma, HR-/HER2+ Ductal Carcinoma, Ductal Triple Negative Breast Cancer (TNBC) and TCGA histological designations Ductal Luminal A, Ductal Luminal B, Ductal HER2-enriched, Ductal Basal-like, Ductal Normal-like. ILC includes MSK-MET histological designations HR+ Lobular Carcinoma and TCGA histological designations Lobular Luminal A, Lobular Luminal B, Lobular HER2-enriched, Lobular Basal-like, Lobular Normal-like. For endometrial cancer, shared cancer types between MSK-MET and TCGA included endometrioid and serous subtypes. For NSCLC, shared cancer types between MSK-MET and TCGA included adenocarcinoma and squamous cell carcinoma. The frequency of WGD was calculated as the number of WGD-positive samples over the total sample count within a self-reported racial group by share histological subtype for each cancer type; testing for association was done via two-sided Pearson’s Chi-squared test across racial groups.

### Concordance between Inferred Genetic Ancestry and Self-reported Race

Inferred genetic ancestry designations (African and European) for each patient of the MSK-MET cohort were sourced from *Arora et al.* and maintained separately. Concordance between inferred genetic ancestry and self-reported race was determined as the proportions of African ancestry patients who self-identified as “Black” and European ancestry patients who self-identified as “white”.

### Correlation of Fractional Ancestry to WGD

Fractional ancestries (AFR and EUR) for all MSK-MET patients regardless of majority inferred genetic ancestry were binned then correlated to the binned rates of WGD. Separately, AFR and EUR fractional ancestry were binned into 5%-sized bins, then within each bin, both the rate of WGD and number of patients were calculated. We then ran a logistic regression between bin number and rate of WGD, weighted by the number of patients, to test for a relationship.

### Logistic Regression of Genomic Aberrations to WGD

In MSK-MET, in order to determine which genomic aberrations correlated with increased rates of WGD, we built multivariate logistic regression models in R for self-reported Black patients only, self-reported white patients only, and all self-reported Black and white patients (i.e., all patients). All frequently occurring genomic aberrations (≥200 patients and ≥2% patients) were included in the models and were coded as binary predictor variables. Using a similar method to Bielski, et al. (2018)^52^, we reduced the models by removal of covariates in two stages: 1) removal of aliased covariates (i.e., those with perfect correlation to another covariate), and 2) recursive removal of the covariate with the highest variance inflation factor (VIF) until all covariates had a VIF≤4 in order to remove multicollinearity. Cancer types were included in the final models. Significance of covariates were tested by Wald test followed by FDR correction via Benjamini & Hochberg’s method^135^.

### Aneuploidy Burden and Chromosomal Arm-Level Differences by Self-Reported Race and WGD Status

Total aneuploidy score, as defined as total amount of chromosome arm gains and losses, were calculated for each patient within MSK-MET and TCGA. Total aneuploidy burden was compared on a pan-cancer level and within cancer types were tested by unpaired t-tests. Chromosomal arm levels frequencies were calculated as the number of samples with chromosome arm aneuploidy (either gain or loss) over the total number of samples with determined arm status (gain, loss, or neutral) for each respective available chromosome arm within each self-report race group on a pan-cancer and cancer type level. Testing for association of specific aneuploidies was done via two-sided Pearson’s Chi-squared test.

### TP53 Analysis

Nonsynonymous mutations in *TP53* were analyzed across racial groups, WGD status, and cancer type. Sample containing at least one nonsynonymous mutation in *TP53* were consider *TP53*-mutant, while the remaining samples were considered *TP53*-WT. The frequency of *TP53* mutation was calculated as the proportion of *TP53*-mutant samples over the total sample count within a racial group at a pan-cancer level and additionally subset by cancer type and/or WGD status; testing for association was done via two-sided Pearson’s Chi-squared test across self-reported racial groups. Variant classification distributions were summarized within each racial group at a pan-cancer level and additionally subset by cancer type; testing for association was done via two-sided Pearson’s Chi-squared test across self-reported racial groups. Location of *TP53* mutations were summarized within each racial group and visually compared across self-reported racial groups.

### CCNE1 Analysis

Gains and losses of CCNE1 were analyzed across racial groups, WGD status, and cancer types. Samples demonstrating amplifications of CCNE1 were considered CCNE1 gain. All remaining samples demonstrating either loss of CCNE1 or no alteration were considered CCNE1 neutral/loss. The frequency of CCNE gains was calculated as the proportion of CCNE1 gained sample over total sample count within a racial group at a pan-cancer level and additional subset by cancer type and/or WGD status; testing for association was done via two-sided Pearson’s Chi-squared test.

### Whole-genome Duplication and Tumor Response and Status

Clinical data including tumor response for first course of treatment and tumor status at the end of the clinical observation period were sourced from TCGA. Tumor responses included for analysis included “Complete Remission/Response”, “Partial Remission/Response”, “Stable Disease”, and “Progressive Disease,” the remaining responses were not included for analysis as these categories represented disputed, uncertain, or missing clinical responses. The frequency of progressive disease was calculated as the proportion of patients with “Progressive Disease” over total number of patients with included tumor responses by WGD status at a pan-cancer and cancer type level. For tumor status, patients with documented “TUMOR_FREE” or “WITH_TUMOR” were included for analysis, remaining patients with missing data were excluded. The frequency of patients with persistent disease at the end of TCGA’s clinical observational period was calculated as the proportion of patients “WITH TUMOR” over total number of patients with documented tumor status by WGD status at a pan-cancer and cancer type level. Testing for association was done via two-sided Pearson’s Chi-squared test.

### Cell Culture

Murine epithelial lung tissue cell line (LA-4; cat. no. ATCC CCL-196) and murine alveolar lung macrophage (MH-S; cat. No. CRL2019) cell line was purchased from and cultured according to American Type Culture Collection (ATCC) instructions. Cells were cultured in TPP cell culture flasks at 37 °C in a 5% CO2 humidified atmosphere until monolayers reached desired confluency. All experiments were performed with cells before the twentieth passage. For long-term live cell experiments, a stage-top incubator that maintains a humidified atmosphere with 5% CO2 and is heated to 37 °C was used. The medium used for culturing of the epithelial LA-4 cells was Ham’s F-12K medium (Gibco) supplemented with 15% FCS (ATCC), 1% P/S (Sigma), 1% NEAA (Gibco), 2 × 10−3 M L-Gln. For alveolar macrophages cell line, MH-S, RPMI 1640 (Gibco) medium supplemented with 10% FCS (ATCC), 1% P/S (Sigma), 2 × 10−3 M L-Gln, and 0.05 × 10−3 M beta mercapthoethanol (Gibco) was used.

### Carcinogen Materials for Exposure Assays

The environmental and engineered particulate matter used in this study are summarized in Table S12. These included four particles produced by fuel combustion (three diesel exhaust samples, one carbon black sample), three engineered carbonaceous particulate matters (graphene oxide, two multiwall carbon nanotubes), two clay samples, and four metal oxides^89,136–148^. TiO_2_ nanotubes were synthesized by Polona Umek^149^. Printex 90 was kindly provided by Evonik, Frankfurt, Germany. NM-401 MWCNT (MWCNTs-NM401-JRCNM04001a) were a kind gift from JRC Nanomaterial Repository. All other materials, except from commercially available DieselN1650, were obtained through the EU project *nanoPASS* from prof. Ulla Vogel (NRCWE, Copenhagen, Denmark).

### Particulate Matter Preparation

Cup horn sonication was employed to disperse particulate matter in a low osmolarity, high pH buffer solution (vehicle: 1mM bicarbonate buffer (100 times diluted bicarbonate buffer), pH of 10) to minimize charge screening of the particulate matters’ active surfaces. To ensure uniform dispersion, particulate matter was resuspended to contain 3 cm^2^ of the particulate matter surface/3uL. The resulting suspensions underwent cup horn sonification in an ice bath for 15 minutes, utilizing five seconds on and five seconds off regimen for a total duration of 30 minutes, at a power setting of 20-30W (amplitude 70) to guarantee optimal dispersion. Prior to microscopy, the volume of particulate matter dispersion containing 10 times larger of the particulate matter surface area of the cell culture well was added in a dropwise manner. The volume of particulate matter applied to cells represented 3% of the final cell media volume.

### Live Cell Microscopy

A combination of fluorophores was used to label structures of interest in cells. We labeled murine epithelial lung tissue cells with CellTracker™ Green CMFDA (CTG, Thermo Fisher (#C2925), excitation peak 492 nm, emission peak 517 nm, 1 μM), CellMask™ Deep Red (CMDR, Thermo Fisher (#C10046), excitation peak 650 nm, emission peak 685 nm, 0.5 μg/mL), and Abberior LIVE 550 Tubulin (Abberior, (#LV550), excitation peak 551 nm, emission peak 573 nm, 100 nM). We labeled murine alveolar lung macrophages with CellTracker™ Orange CMRA dye (CTO, Thermo Fisher (#C34551), excitation peak 548 nm, emission peak 576 nm, 1 μM). It is noteworthy that CTG and CTO fluorophores were added one day prior to microscopy, while the other fluorophores were added immediately before imaging. Additionally, only CTG and CTO were washed with PBS, whereas the other labels were not. Live cell microscopy was performed using the STED microscope by Abberior Instruments in confocal mode. An inverted microscope body (Olympus IX83) was equipped with a stage top incubator (Okolab H301-MIN), which maintains atmosphere with the 37°C, 5% CO2, and at least 95% humidity to enable long term imaging of living cells. Images were captured by 20x magnification and 0.8 numerical aperture (NA) lens. The microscope system incorporates four pulsed laser sources with a pulse duration of 120 ps and a maximum power of 50 µW at the sample plane. Four avalanche photodiode (APD) detectors are utilized for signal detection. We detected particulate matter in the label-free, backscatter detection mode, utilizing the 488 / 488 ± 5 nm excitation / detection.

### Carcinogen Exposure Assays

Cells labeled with live cell compatible fluorophores were exposed to particulate matter for a total of 24 hours as detailed in *Kokot et al*.^89^. Cytotracker was added one day prior to microscopy and removed prior to microscopy. The remaining fluorophores were added immediately prior to imaging. Murine lung epithelial cells in monoculture or in coculture with murine macrophages were exposed to individual particulate matters immediately prior to imaging. For each particulate matter, 3-4 biological replicates were performed (cells with the next (+1) passage number, seeded and measured 3-4 days later), each with 1-3 technical replicates (cells with the same passage number, but seeded in a neighboring well and measured in parallel on the same day). Control samples are cells labeled with all fluorophores exposed to 3 μL of vehicle buffer without particulate matter (1 mM (100 x diluted) bicarbonate buffer only), representing 3% of the total cell medium volume. 24-hour time-lapse fluorescence and scattering microscopy images were quantified utilizing standard quantification algorithms of the Infinite platform (version 42), written in Python and Mathematica, to derive: 1) cell proliferation (number of cells), 2) binucleate cell formation (fraction of bi- and multinucleate epithelial cells). Rates of proliferation and binucleation changes were averaged within all the available biological and/or technical replicates. Changes in cellular responses to carcinogen exposures were tested by pairwise two-tailed t-tests to reference control.

### Mutagen Signature Attributions

In order to extract mutagen signature attributions from TCGA NSCLC samples, the ‘signatureFit_pipelin’ of signature.tools.lib was run using the Mutagen53 catalogue from Kucab, et al. (2019), with the settings: genome.v = “hg19”, randomSeed = 6206, fit_method = “Fit”, threshold_p.value = 0.05, optimisation_method = “KLD”, useBootstrap = FALSE, exposureFilterType = “fixedThreshold”, threshold_percent = 5, threshold_nmuts = 10, multiStepMode = “errorReduction”, and minErrorReductionPerc = 15. Results were then saved via ‘plotFitResults’ from the same package. These results were then normalized across samples by taking the number of assignments per mutagen signature in a sample divided by the total assignments for that sample. These normalized assignments were then summed within the mutagen classes as defined by Kucab, et al. (2019). We then compared these mutagen class assignments across self-reported racial groups using two-sided Welch’s t-tests followed by false discovery rate correction using Benjamini-Hochberg’s method.

### Survival Analysis

Survival analysis was performed in Python, using the packages: lifelines, matplotlib, and pandas. Significance was tested via logrank test.

### Clinical Correlates of WGD and Patient Race

Equivalence of ages at key clinical event times (diagnosis, death, sequencing, and surgical procedure) were compared across self-reported racial groups and WGD status via Wilcoxon rank-sum test. All ages except for diagnosis were directly reported by MSK-MET, while age at diagnosis was inferred based on survival status, whereby dead patients’ age at diagnosis was calculated as overall survival in months, while for living patients’ age at diagnosis was calculated from age at last contact minus overall survival in months. Microsatellite instability status was determined using designated Stable and Instable determinations; the frequency of microsatellite instability (MSI-H) was determined as the number of Instable samples divided by the total number of Stable and Instable samples within a self-reported racial group or WGD status; testing for association was done via two-sided Pearson’s Chi-squared test. Staging information for MSK-MET dataset remained incomplete. The frequency of metastasis location was determined by the proportion of samples with recorded metastasis divided by the total number of samples from a self-reported racial group or WGD status; testing for association was done via two-sided Pearson’s Chi-squared test. For those metastases occurring in only a subset of patients (e.g., Female Genital and Male Genital), only samples contained within the same subset were considered in our calculations.

### Data Visualization

Scientific illustrations were assembled using BioRender. Graphs and scatterplots were generated using Graphpad Prism.

## Data Availability

All data referenced in the manuscript are publicly available.

## ACKNOWLEDGMENTS

We thank John Kunstman, Sajid Khan, Jun Lu, Marc Vittoria, Kanika Arora, Elizabeth Godfrey, Danielle Heller, Daniel Kerekes, Marcella Nunez-Smith, and members of the Sheltzer Lab for helpful feedback on this manuscript. The authors kindly thank JRC Nanomaterials Repository for providing various the Multiwall carbon nanotubes (JRCNM04001a, former code NM-401), and Nanoclay Bentonite (JRCNM0600a, former code NM-600) used in this study from the JRC Nanomaterials Repository. This research was funded by the Slovenian Research and InnovSation Agency (program P1-0060) and European Union (nanoPASS, 101092741). Research in the Sheltzer Lab is supported by NIH grants R01CA237652 and R01CA276666, Department of Defense grant W81XWH-20-1-068, an American Cancer Society Research Scholar Grant, a Breast Cancer Alliance Young Investigator Award, a sponsored research agreement from Ono Pharmaceuticals, and a sponsored research agreement from Meliora Therapeutics.

## SUPPLEMENTAL FIGURE LEGENDS

**Figure S1.**
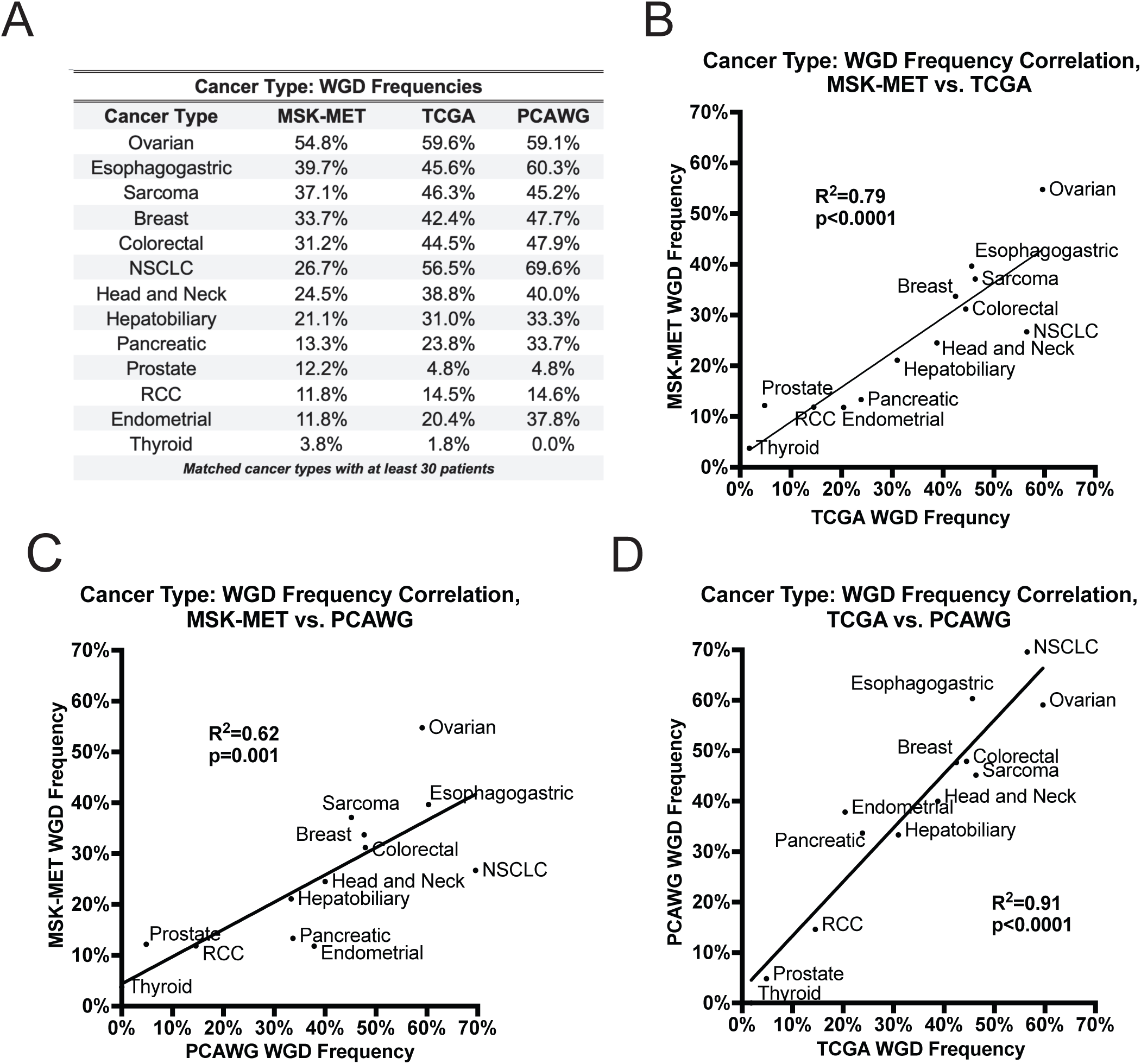
Frequency of WGD events by cancer type. **(A)** A table showing the frequency of WGD events in different cancer types across the MSK-MET, TCGA, and PCAWG patient cohorts. **(B)** A scatterplot showing the correlation between the frequency of WGD events within cancer types between the TCGA and MSK-MET datasets. **(C)** A scatterplot showing the correlation between the frequency of WGD events within cancer types between the MSK-MET and PCAWG datasets. **(D)** A scatterplot showing the correlation between the frequency of WGD events within cancer types between the TCGA and PCAWG datasets. Acronyms: renal cell carcinoma (RCC). Correlation was conducted via Pearson correlation (B-D).

**Figure S2.**
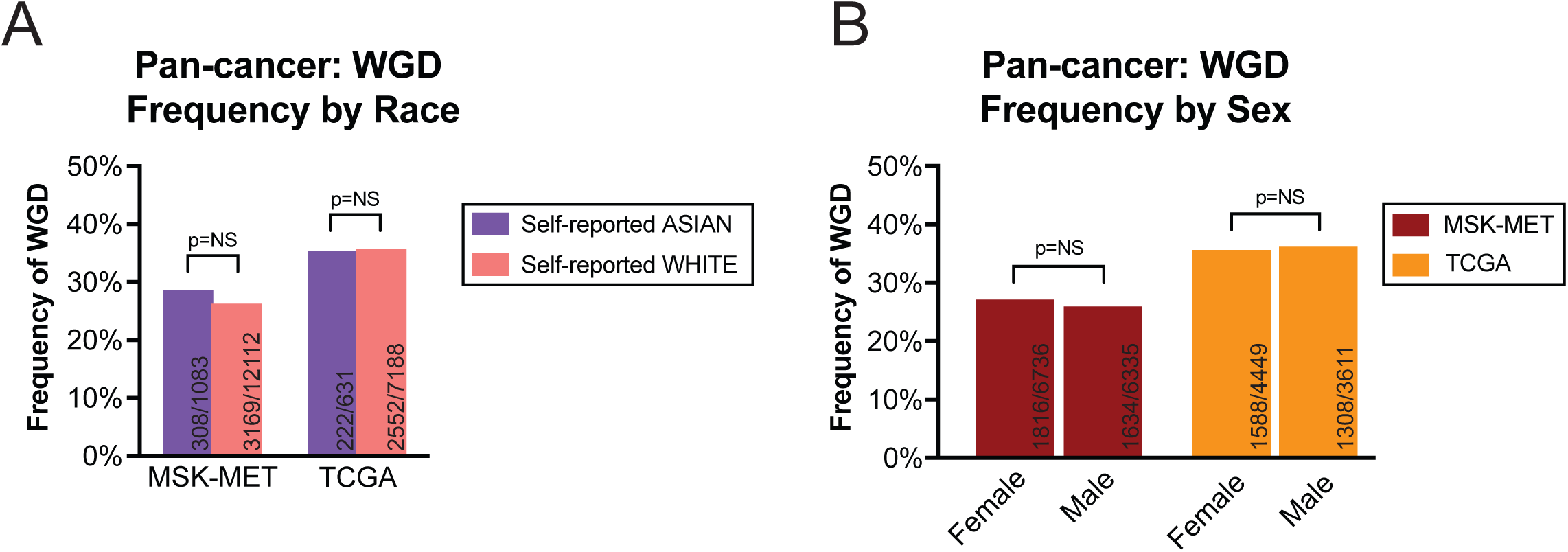
WGD status among self-reported Asian patients and patient sex. **(A)** A bar graph displaying the frequency of WGD events among tumors from self-reported Asian and white cancer patients. **(B)** A bar graph displaying the frequency of WGD events among tumors from female and male cancer patients. Statistical testing was performed via two-tailed Pearson’s Chi-squared test (A-B). Statistical significance: NS p ≥ 0.05, * p < 0.05, ** p < 0.01, *** p < 0.001, **** p < 0.0001.

**Figure S3.**
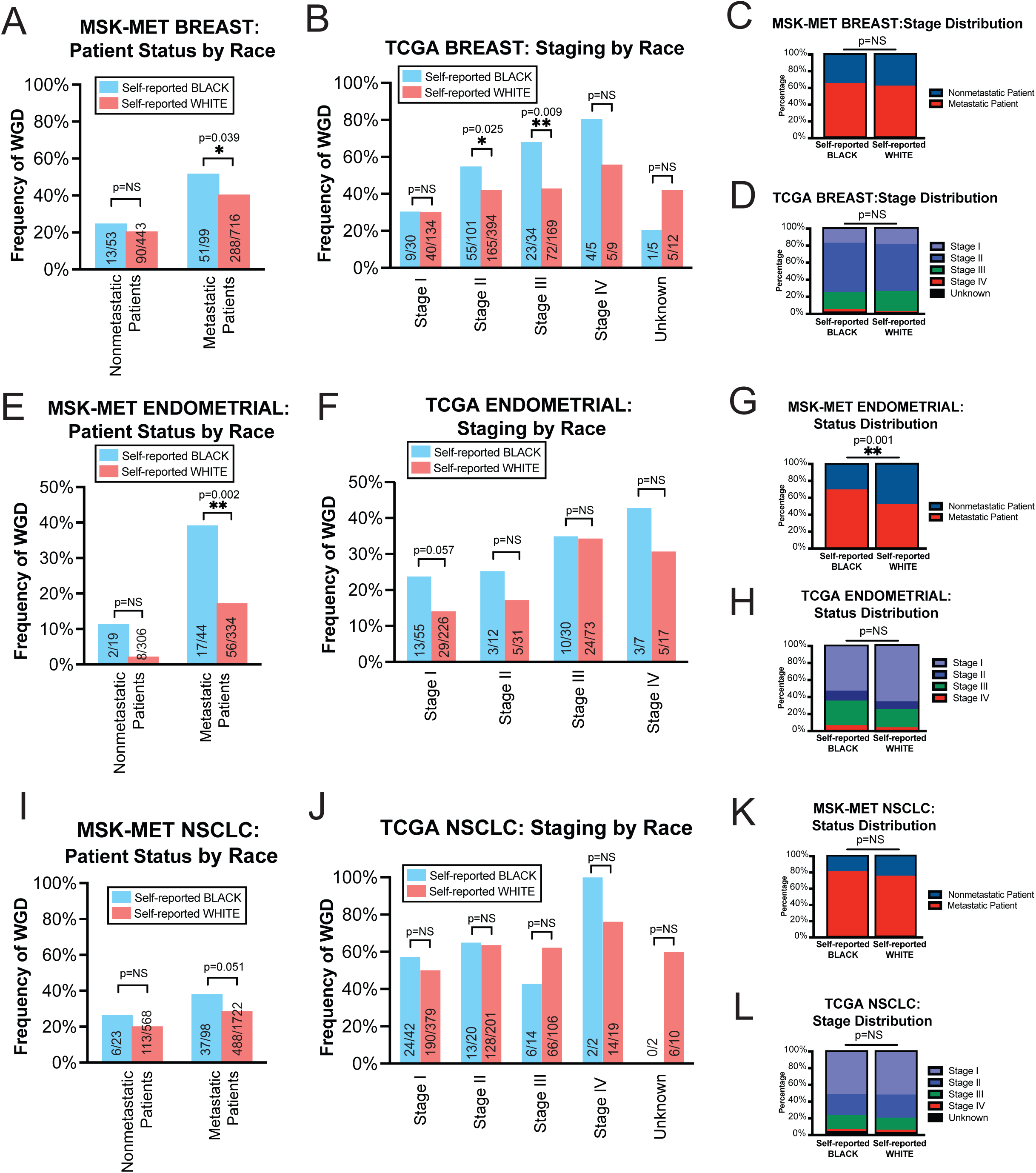
WGD status separated based on metastatic status and tumor stage. **(A)** A bar graph displaying the frequency of WGD events among tumors from self-reported Black and white patients stratified by metastatic status in the MSK-MET breast cancer cohort. **(B)** A bar graph displaying the frequency of WGD events among tumors from self-reported Black and white patients stratified by pathological stage in TCGA breast cancer cohort. **(C)** A bar graph displaying the distribution of metastatic status by self-reported race within the MSK-MET breast cancer cohort. **(D)** A bar graph displaying the distribution of pathological stage by self-reported race within the TCGA breast cancer cohort. **(E)** A bar graph displaying the frequency of WGD events among tumors from self-reported Black and white patients stratified by metastatic status in the MSK-MET endometrial cancer cohort. **(F)** A bar graph displaying the frequency of WGD events among tumors from self-reported Black and white patients stratified by clinical stage in TCGA endometrial cancer cohort. **(G)** A bar graph displaying the distribution of metastatic status by self-reported race within the MSK-MET endometrial cancer cohort. **(H)** A bar graph displaying the distribution of clinical stage by self-reported race within the TCGA endometrial cancer cohort. **(I)** A bar graph displaying the frequency of WGD events among tumors from self-reported Black and white patients stratified by metastatic status in the MSK-MET NSCLC cancer cohort. **(J)** A bar graph displaying the frequency of WGD events among tumors from self-reported Black and white patients stratified by pathological stage in TCGA NSCLC cancer cohort. **(K)** A bar graph displaying the distribution of metastatic status by self-reported race within the MSK-MET NSCLC cancer cohort. **(L)** A bar graph displaying the distribution of pathological stage by self-reported race within the TCGA NSCLC cancer cohort. Statistical testing was performed via Fisher’s exact test (A, B, E, F, I, J) and Pearson’s Chi-squared test (C, D, G, H, K, L). Statistical significance: NS p ≥ 0.05, * p < 0.05, ** p < 0.01, *** p < 0.001, **** p < 0.0001.

**Figure S4.**
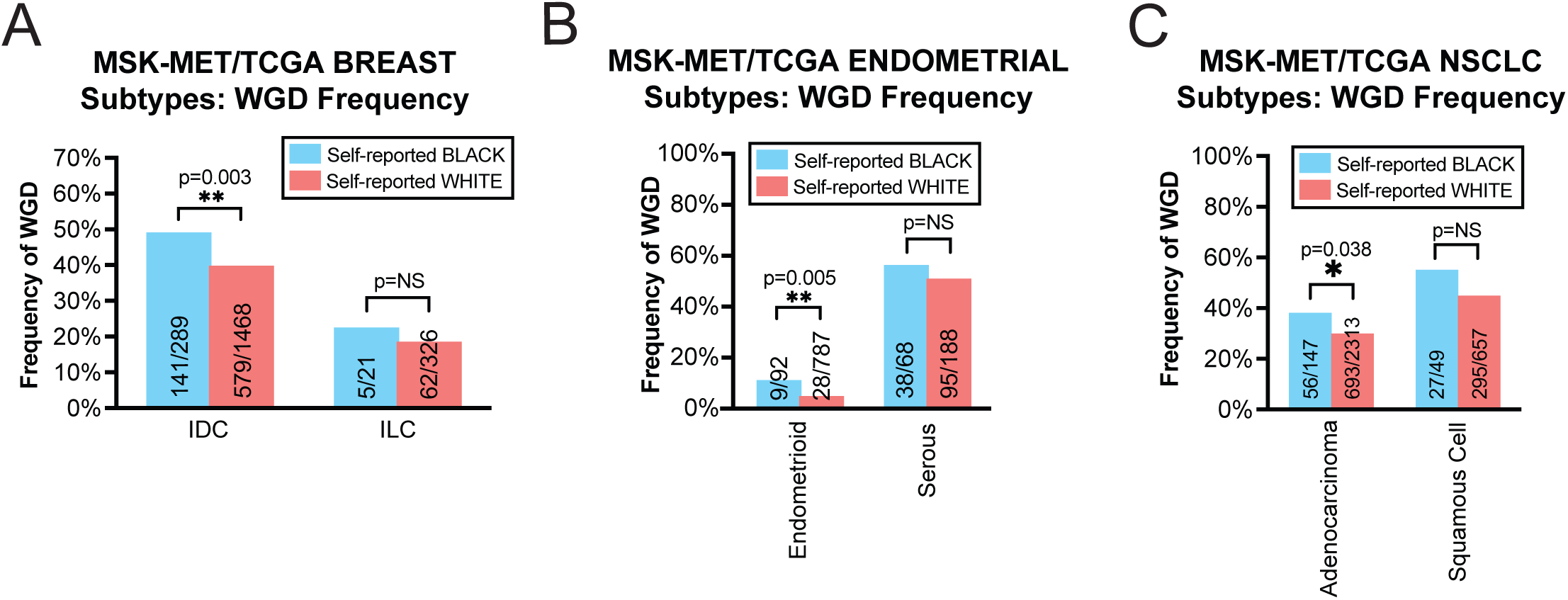
WGD status separated based on histological subtypes. **(A)** A bar graph displaying the frequency of WGD events among tumors from self-reported Black and white patients from shared breast cancer histological subtypes in MSK-MET and TCGA cohorts. **(B)** A bar graph displaying the frequency of WGD events among tumors from self-reported Black and white patients from shared endometrial cancer histological subtypes in MSK-MET and TCGA cohorts. **(C)** A bar graph displaying the frequency of WGD events among tumors from self-reported Black and white patients from shared endometrial cancer histological subtypes in MSK-MET and TCGA cohorts. Acronyms: Invasive/Infiltrating Ductal Carcinoma (IDC), Invasive/Infiltrating Carcinoma (ILC). Statistical testing was performed via Pearson’s Chi-squared test (A-C). Statistical significance: NS p ≥ 0.05, * p < 0.05, ** p < 0.01, *** p < 0.001, **** p < 0.0001.

**Figure S5.**
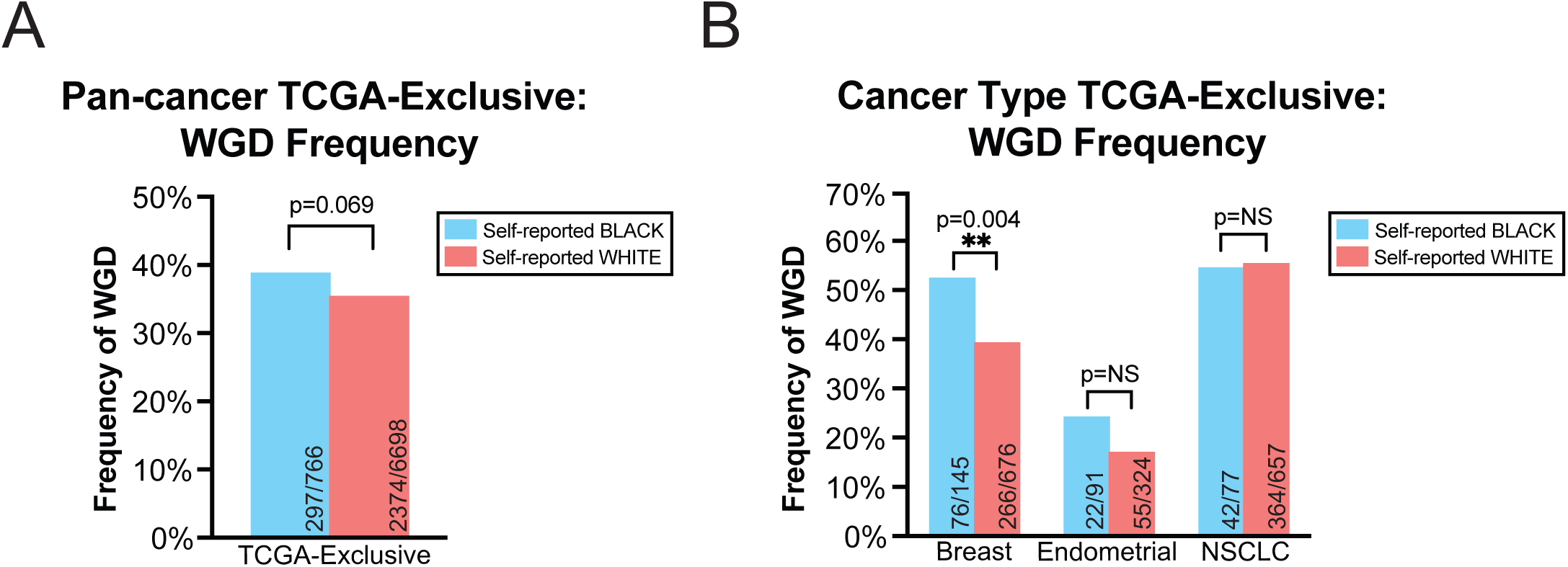
Frequency of WGD in TCGA-exclusive cohort. **(A)** A bar graph displaying the frequency of WGD events in self-reported Black and white patients of the TCGA-exclusive pan-cancer cohort. **(B)** A bar graph displaying the frequency of WGD events in self-reported Black and white patients of the TCGA-exclusive in breast cancer, endometrial cancer, or NSCLC. Statistical testing was performed via two-tailed Pearson’s Chi-squared test (A-B). Statistical significance: NS p ≥ 0.05, * p < 0.05, ** p < 0.01, *** p < 0.001, **** p < 0.0001.

**Figure S6.**
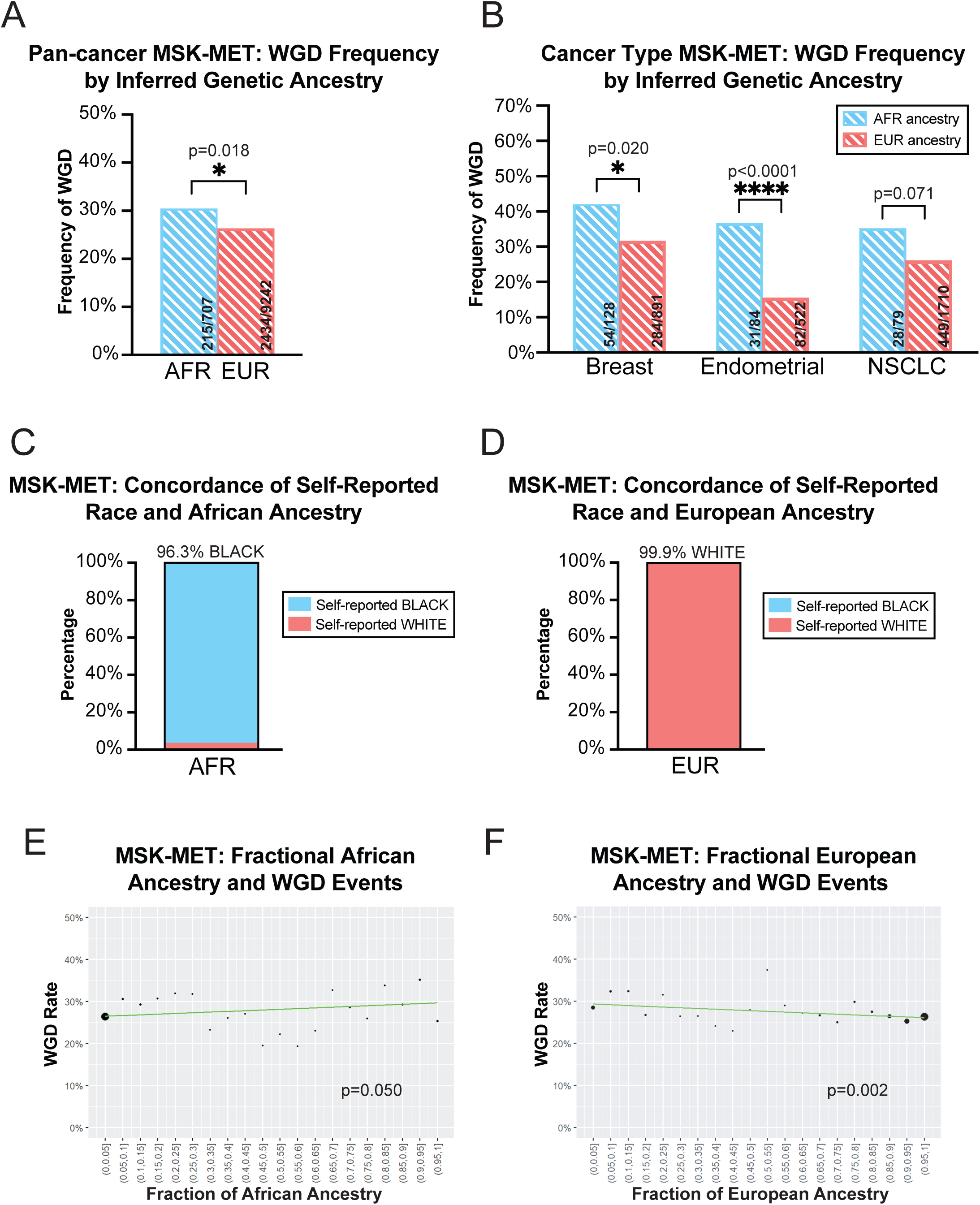
Analysis of WGD status based on inferred genetic ancestry. **(A)** A bar graph displaying the frequency of WGD events among tumors from cancer patients with inferred African ancestry (AFR) and European ancestry (EUR). **(B)** A bar graph displaying the frequency of WGD events among patients with either breast cancer, endometrial cancer, or NSCLC based on inferred genetic ancestry. **(C)** A stacked bar graph displaying the proportion of self-reported Black patients within patients with AFR inferred genetic ancestry. **(D)** A stacked bar graph displaying the proportion of self-reported white patients within patients with EUR inferred genetic ancestry. **(E)** Correlation of fractional African ancestry to rates of WGD for all MSK-MET patients regardless of majority inferred genetic ancestry. Point sizes correspond to the number of patients in the bin. **(F)** Correlation of fractional European ancestry to rates of WGD for all MSK-MET patients regardless of majority inferred genetic ancestry. Point sizes correspond to the number of patients in the bin. Statistical testing was performed via two-tailed Pearson’s Chi-squared test (A-B). Statistical significance: NS p ≥ 0.05, * p < 0.05, ** p < 0.01, *** p < 0.001, **** p < 0.0001.

**Figure S7.**
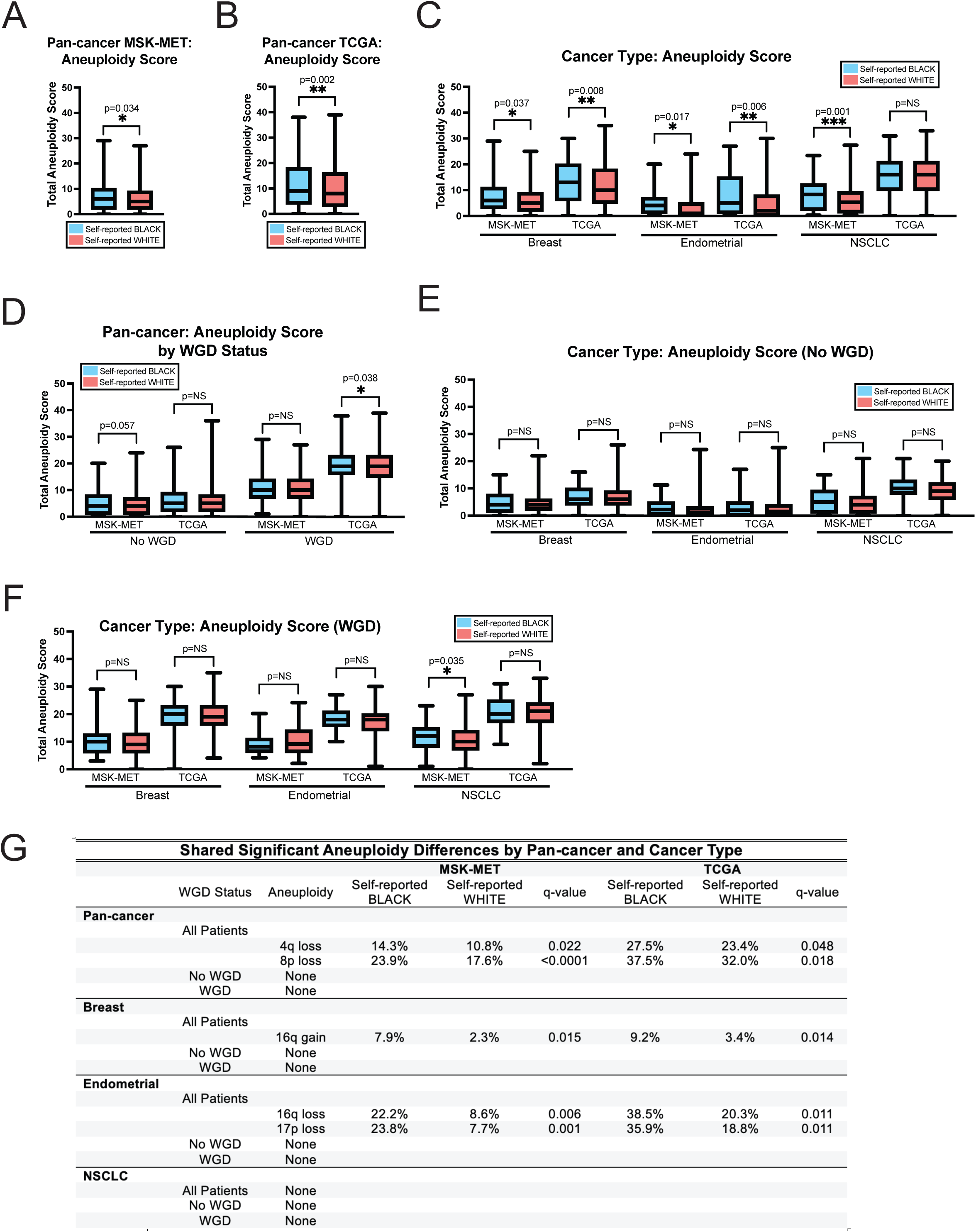
Aneuploidy burden and aneuploidy differences by self-reported race and WGD status. Boxplots displaying the total aneuploidy score of self-reported Black and white patients in (**A)** MSK-MET **(B)** TCGA. **(C)** A boxplot displaying the total aneuploidy score of self-reported Black and white patients in MSK-MET and TCGA cohorts in breast cancer, endometrial cancer, and NSCLC. **(D)** A boxplot displaying the total aneuploidy score of self-reported Black and white patients in MSK-MET and TCGA cohorts by WGD status. **(E)** A boxplot displaying the total aneuploidy score of self-reported Black and white WGD-negative patients in MSK-MET and TCGA breast cancer, endometrial cancer, and NSCLC cohorts. **(F)** A boxplot displaying the total aneuploidy score of self-reported Black and white WGD-positive patients in MSK-MET and TCGA breast cancer, endometrial cancer, and NSCLC cohorts. **(G)** A table of shared significant chromosome arm-level aneuploidy differences within self-reported Black and white patients across MSK-MET and TCGA cohorts. Statistical testing was performed via unpaired t-test (A-F) and Pearson’s Chi-squared test followed by Benjamini-Hochberg correction^135^. (G). Statistical significance: NS p ≥ 0.05, * p < 0.05, ** p < 0.01, *** p < 0.001, **** p < 0.0001.

**Figure S8.**
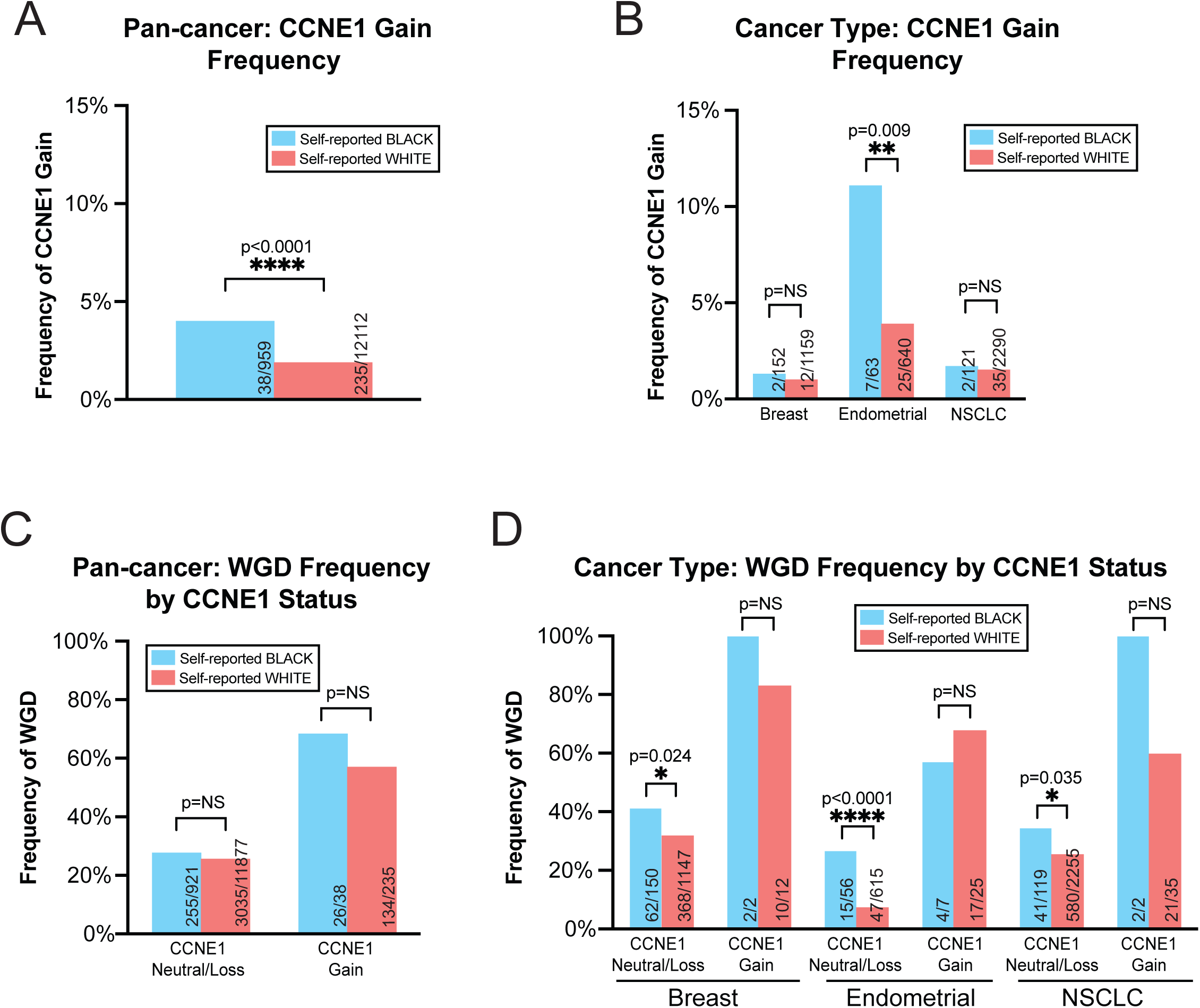
CCNE1 status by self-reported race and WGD status. **(A)** A bar graph displaying the frequency of CCNE1 gains in self-reported Black and white patients. **(B)** A bar graph displaying the frequency of CCNE1 gains in self-reported Black and white patients in breast cancer, endometrial cancer, NSCLC. **(C)** A bar graph displaying the frequency of WGD events in self-reported Black and white patients by CCNE1 status. **(D)** A bar graph displaying the frequency of WGD events in self-reported Black and white patients by CCNE1 status in breast cancer, endometrial cancer, NSCLC. Statistical testing was performed via two-tailed Pearson’s Chi-squared test (A-D). Statistical significance: NS p ≥ 0.05, * p < 0.05, ** p < 0.01, *** p < 0.001, **** p < 0.0001.

**Figure S9.**
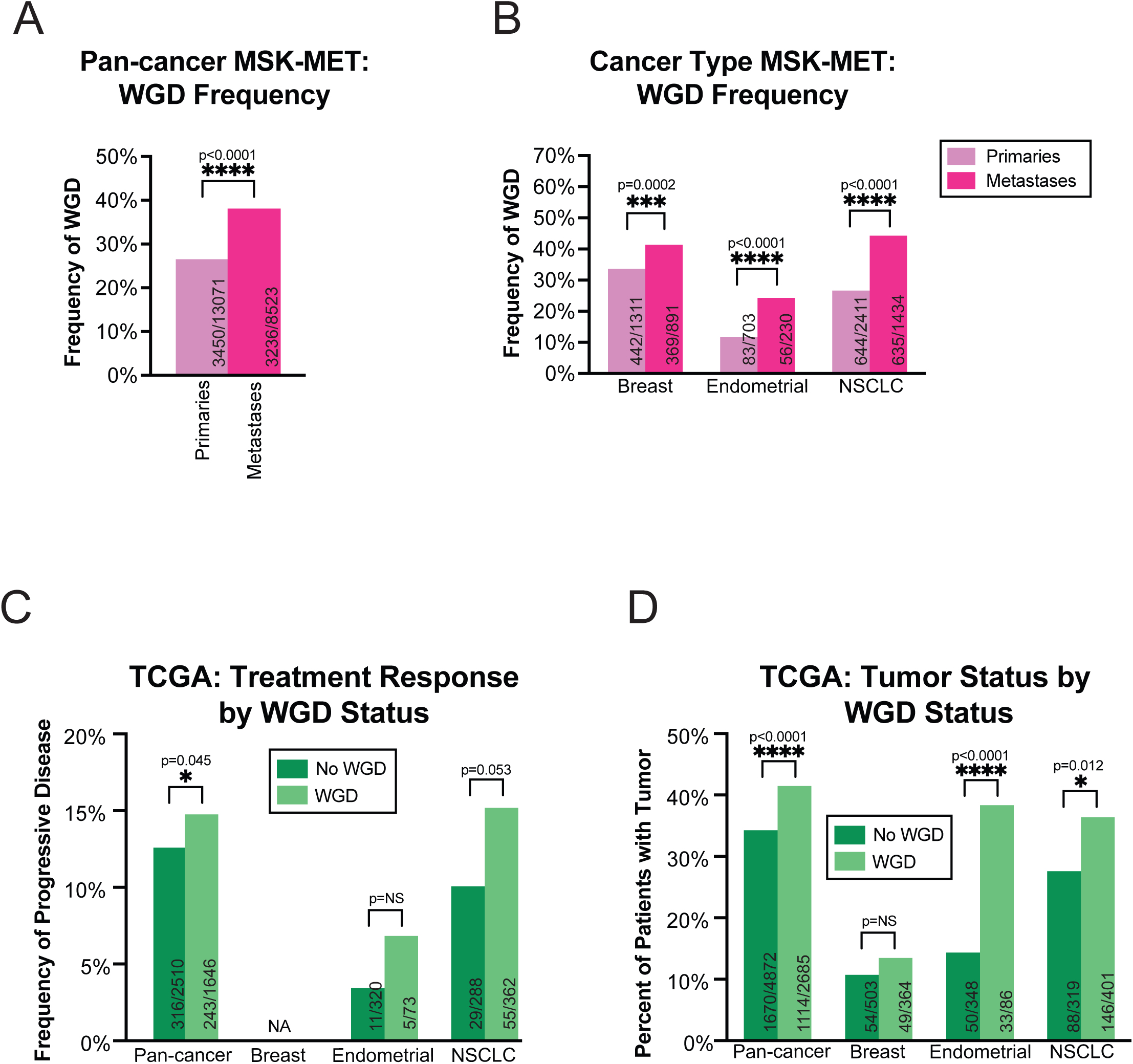
WGD by sample type, treatment response, and observational outcome. **(A)** A bar graph displaying the frequency of WGD events in primaries and metastases. **(B)** A bar graph displaying the frequency of WGD events in primaries and metastases in breast cancer, endometrial cancer, and NSCLC. **(C)** A bar graph displaying the frequency of progressive disease of WGD-negative and WGD-positive tumors by pan-cancer analysis and cancer type (breast cancer, endometrial cancer, NSCLC). **(D)** A bar graph displaying the frequency of tumor presence at the end of TCGA observational cohort of WGD-negative and WGD-positive tumors by pan-cancer analysis and cancer type (breast cancer, endometrial cancer, NSCLC). Statistical testing was performed via two-tailed Pearson’s Chi-squared test (A-D). Statistical significance: NS p ≥ 0.05, * p < 0.05, ** p < 0.01, *** p < 0.001, **** p < 0.0001.

**Figure S10.**
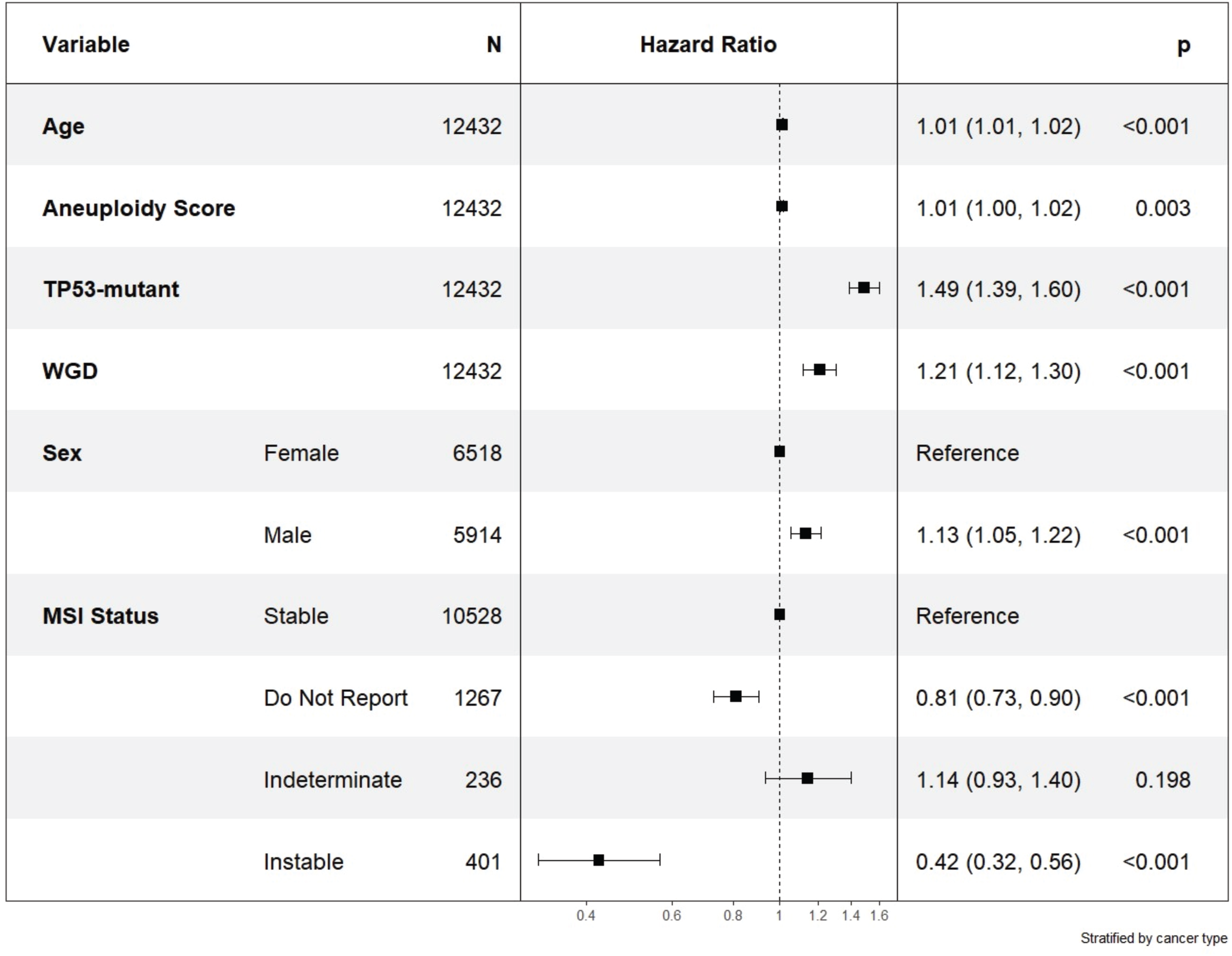
Cox Proportional Hazards analysis of WGD events. A forest plot showing the results of a multivariate, stratified Cox proportional hazard model of overall survival for Black and white MSK-MET patients. The Cox proportional hazard model was stratified by cancer type and constructed with known clinical confounders (age, sex, and MSI status), critical mutations (TP53 mutation), and aneuploidy covariates (aneuploidy score and WGD status). Aneuploidy score is the sum of chromosomal arm-level aneuploidies. Statistical testing was performed via Wald test. Hazard ratio estimates are listed along with their 95% confidence intervals.

**Figure S11.**
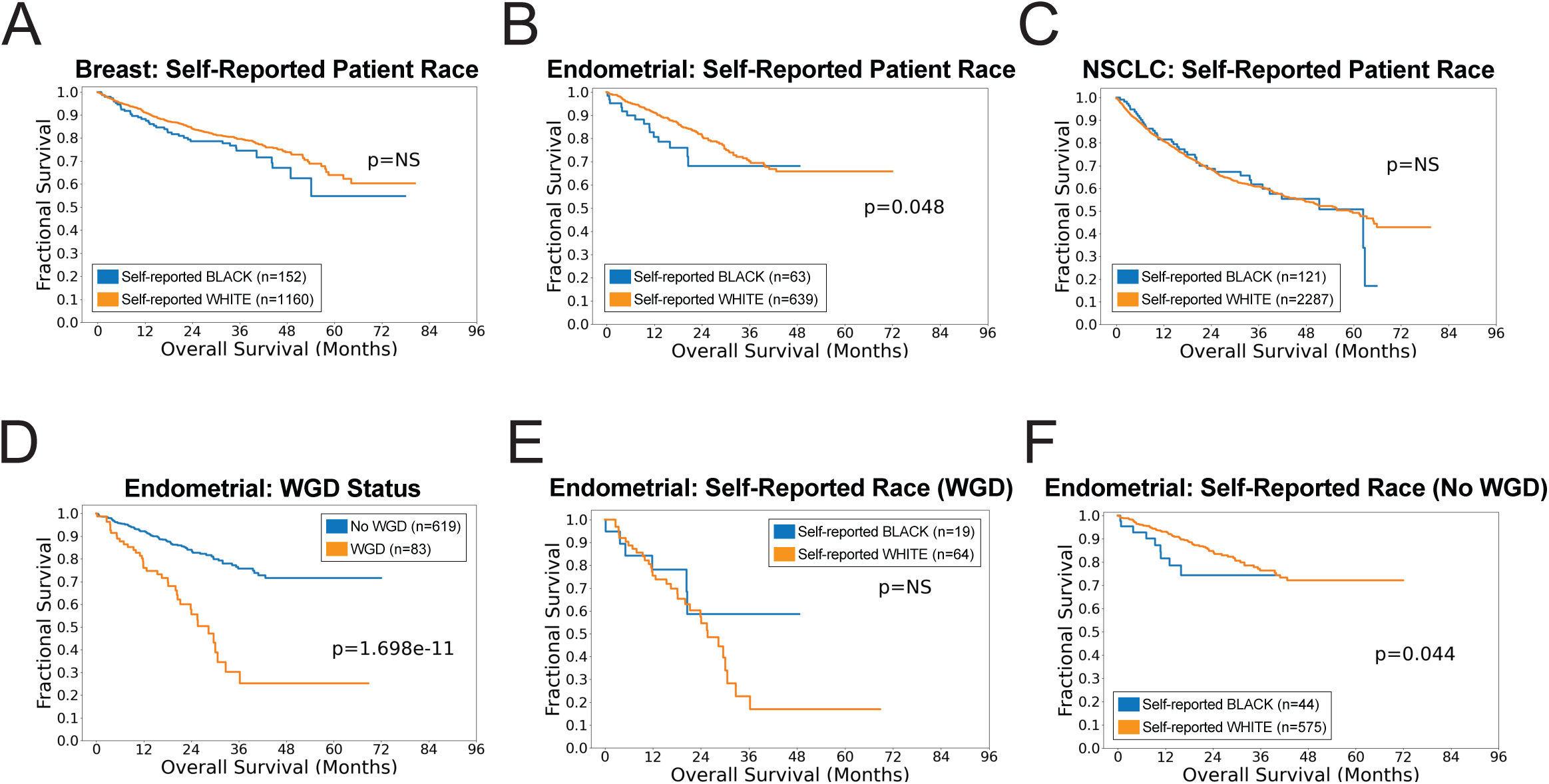
Survival analysis by cancer type in self-reported Black and white cancer patients. **(A-C)** A Kaplan-Meier plot displaying survival of self-reported Black and white patients with **(A)** breast cancer, **(B)** endometrial cancer, and **(C)** NSCLC. **(D)** A Kaplan-Meier plot displaying survival of endometrial cancer patients based on WGD status. **(E)** A Kaplan-Meier plot displaying the survival of WGD-positive self-reported Black and white endometrial cancer patients. **(F)** A Kaplan-Meier plot displaying the survival of WGD-negative self-reported Black and white endometrial cancer patients. Statistical testing was performed via logrank test (A-F).

**Figure S12.**
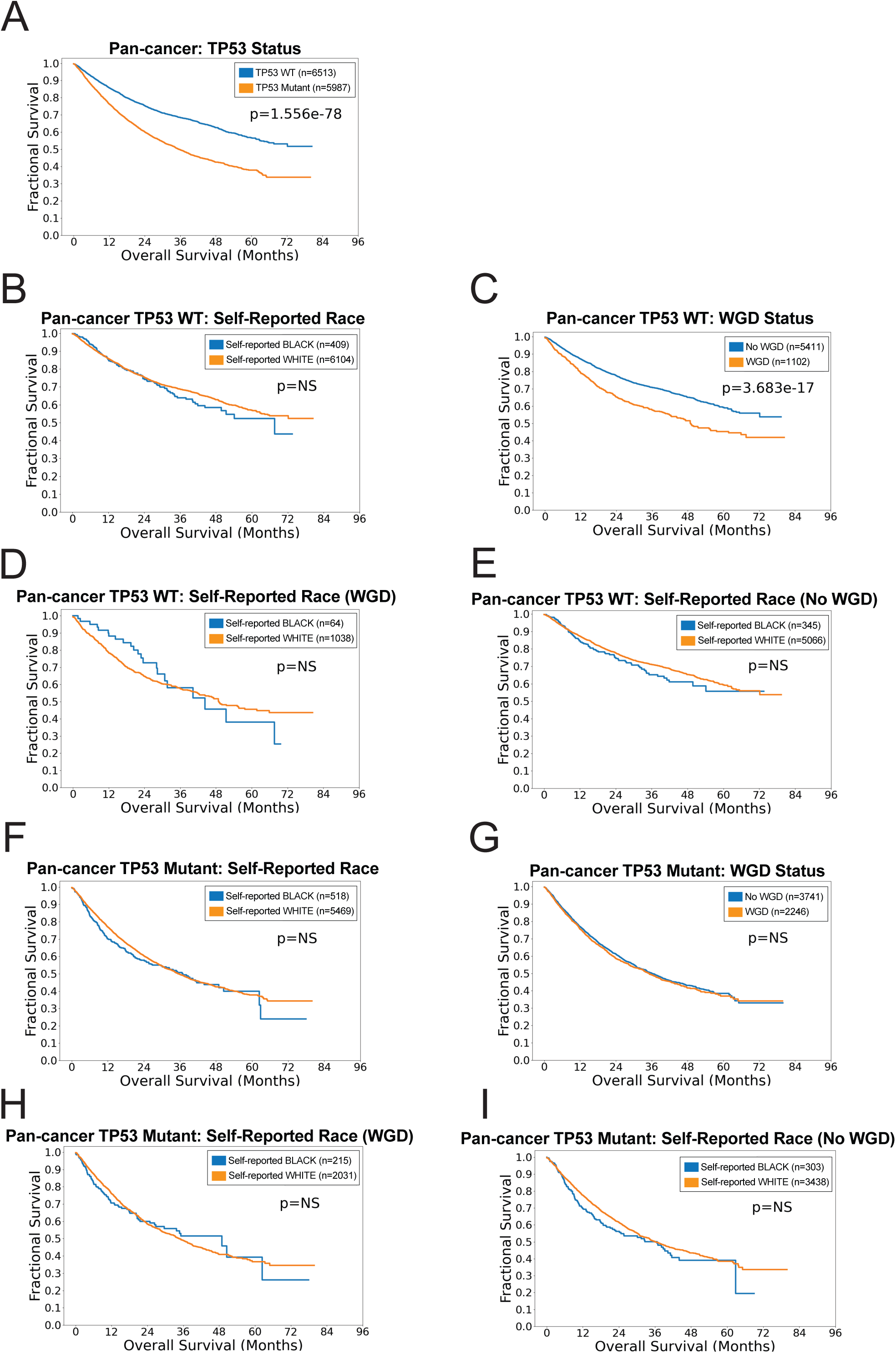
Association between WGD events, *TP53* status, patient self-reported race, and patient survival. **(A)** A Kaplan-Meier plot displaying the survival of cancer patients based on *TP53* mutation status. **(B)** A Kaplan-Meier plot displaying the survival of *TP53*-WT self-reported Black and white cancer patients. **(C)** A Kaplan-Meier plot displaying the survival of *TP53*-WT cancer patients based on WGD status. **(D)** A Kaplan-Meier plot displaying the survival of *TP53*-WT, WGD-positive self-reported Black and white cancer patients. **(E)** A Kaplan-Meier plot displaying the survival of *TP53*-WT, WGD-negative self-reported Black and white cancer patients. **(F)** A Kaplan-Meier plot displaying the survival of *TP53*-mutant self-reported Black and white cancer patients. **(G)** A Kaplan-Meier plot displaying the survival of *TP53*-mutant cancer patients based on WGD status. **(H)** A Kaplan-Meier plot displaying the survival of *TP53*-mutant, WGD-positive self-reported Black and white cancer patients. **(I)** A Kaplan-Meier plot displaying the survival of *TP53*-mutant, WGD-negative self-reported Black and white cancer patients. Statistical testing was performed via logrank test (A-I).

**Figure S13.**
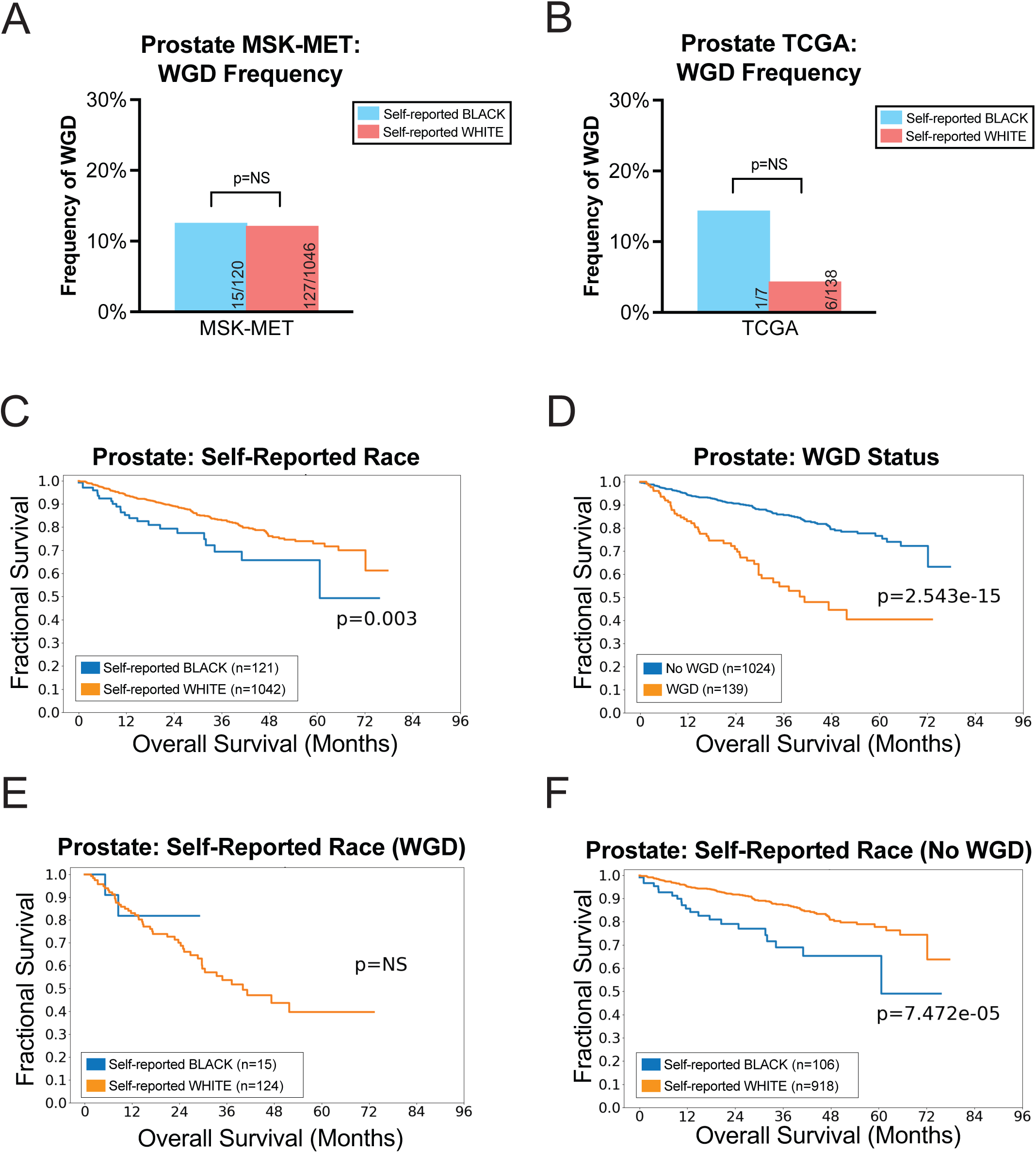
WGD frequency and survival analysis in prostate cancer. **(A)** A bar graph displaying the frequency of WGD events in self-reported Black and white prostate cancer patients in MSK-MET. **(B)** A bar graph displaying the frequency of WGD events in self-reported Black and white prostate cancer patients in TCGA. **(C)** A Kaplan-Meier plot displaying survival of prostate cancer patients by self-reported race. **(C)** A Kaplan-Meier plot displaying survival of prostate cancer patients by WGD status. **(E)** A Kaplan-Meier plot displaying the survival of WGD-positive prostate cancer patients by self-reported race. **(F)** A Kaplan-Meier plot displaying the survival of WGD-negative prostate cancer patients by self-reported race. Statistical testing was performed via two-tailed Pearson’s Chi-squared test (A-B) and logrank test (C-F). Statistical significance: NS p ≥ 0.05, * p < 0.05, ** p < 0.01, *** p < 0.001, **** p < 0.0001.

**Figure S14.**
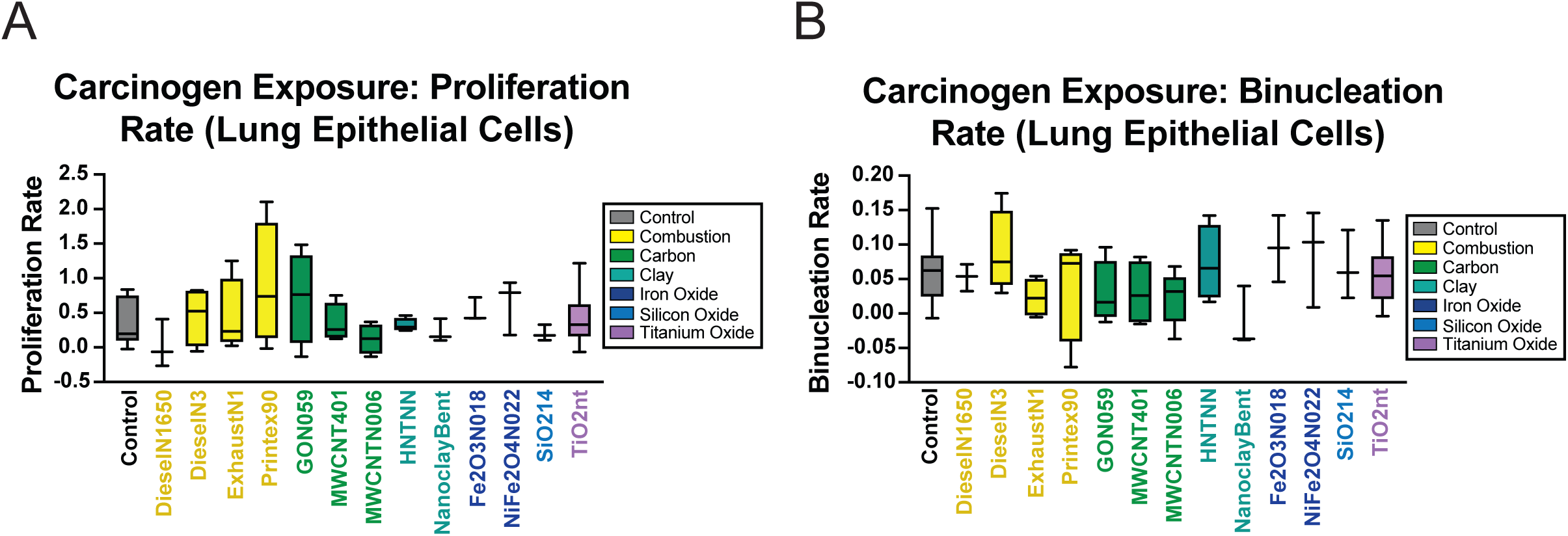
Carcinogen exposure effects in monocultures of lung epithelial cells. **(A)** A boxplot displaying proliferation rate of monoculture lung epithelial cells after control and carcinogen exposure. **(B)** A boxplot displaying binucleation rate of monoculture lung epithelial cells and carcinogen exposure. Statistical testing was performed via pairwise two-tailed t-tests to reference control. Statistical significance: NS p ≥ 0.05, * p < 0.05, ** p < 0.01, *** p < 0.001, **** p < 0.0001.

## References

1. Caraballo, C. et al. Temporal Trends in Racial and Ethnic Disparities in Multimorbidity Prevalence in the United States, 1999-2018. Am. J. Med. 135, 1083–1092.e14 (2022).

2. National Academies of Sciences, E. et al. The State of Health Disparities in the United States. In Communities in Action: Pathways to Health Equity (National Academies Press (US), 2017).

3. Rich, J. A. Primary Care for Young African American Men. J. Am. Coll. Health 49, 183–186 (2001).

4. Williams, D. R. & Mohammed, S. A. Discrimination and racial disparities in health: evidence and needed research. J. Behav. Med. 32, 20–47 (2009).

5. Heller, D. R., Nicolson, N. G., Ahuja, N., Khan, S. & Kunstman, J. W. Association of Treatment Inequity and Ancestry With Pancreatic Ductal Adenocarcinoma Survival. JAMA Surg. 155, e195047 (2020).

6. Sherman, M. E. & Devesa, S. S. Analysis of racial differences in incidence, survival, and mortality for malignant tumors of the uterine corpus. Cancer 98, 176–186 (2003).

7. Megwalu, U. C. & Ma, Y. Racial Disparities in Oropharyngeal Cancer Stage at Diagnosis. Anticancer Res. 37, 835–839 (2017).

8. O’Keefe, E. B., Meltzer, J. P. & Bethea, T. N. Health disparities and cancer: racial disparities in cancer mortality in the United States, 2000-2010. Front. Public Health 3, 51 (2015).

9. Aizer, A. A. et al. Lack of reduction in racial disparities in cancer-specific mortality over a 20-year period. Cancer 120, 1532–1539 (2014).

10. Islami, F. et al. American Cancer Society’s report on the status of cancer disparities in the United States, 2021. CA. Cancer J. Clin. 72, 112–143 (2022).

11. Yedjou, C. G. et al. Health and Racial Disparity in Breast Cancer. Adv. Exp. Med. Biol. 1152, 31–49 (2019).

12. Ryan, B. M. Lung cancer health disparities. Carcinogenesis 39, 741–751 (2018).

13. Siegel, R. L., Miller, K. D., Fuchs, H. E. & Jemal, A. Cancer statistics, 2022. CA. Cancer J. Clin. 72, 7– 33 (2022).

14. D’Arcy, M. et al. Race-associated biological differences among Luminal A breast tumors. Breast Cancer Res. Treat. 152, 437–448 (2015).

15. Beebe-Dimmer, J. L. et al. Racial Differences in Risk of Prostate Cancer Associated With Metabolic Syndrome. Urology 74, 185–190 (2009).

16. Dess, R. T. et al. Association of Black Race With Prostate Cancer–Specific and Other-Cause Mortality. JAMA Oncol. 5, 975 (2019).

17. Olson, S. H. et al. The Impact of Race and Comorbidity on Survival in Endometrial Cancer. Cancer Epidemiol. Biomarkers Prev. 21, 753–760 (2012).

18. Cote, M. L., Ruterbusch, J. J., Olson, S. H., Lu, K. & Ali-Fehmi, R. The Growing Burden of Endometrial Cancer: A Major Racial Disparity Affecting Black Women. Cancer Epidemiol. Biomarkers Prev. 24, 1407– 1415 (2015).

19. Chornokur, G., Dalton, K., Borysova, M. & Kumar, N. Disparities at Presentation, Diagnosis, Treatment and Survival in African American Men, Affected by Prostate Cancer. The Prostate 71, 985–997 (2011).

20. Park, E. R., Japuntich, S. J., Traeger, L., Cannon, S. & Pajolek, H. Disparities Between Blacks and Whites in Tobacco and Lung Cancer Treatment. The Oncologist 16, 1428–1434 (2011).

21. Williams, D. R. & Rucker, T. D. Understanding and Addressing Racial Disparities in Health Care. Health Care Financ. Rev. 21, 75–90 (2000).

22. Coughlin, S. S., King, J., Richards, T. B. & Ekwueme, D. U. Cervical Cancer Screening among Women in Metropolitan Areas of the United States by Individual-Level and Area-Based Measures of Socioeconomic Status, 2000 to 2002. Cancer Epidemiol. Biomarkers Prev. 15, 2154–2159 (2006).

23. Stormacq, C., Van den Broucke, S. & Wosinski, J. Does health literacy mediate the relationship between socioeconomic status and health disparities? Integrative review. Health Promot. Int. 34, e1–e17 (2019).

24. Beech, B. M., Ford, C., Thorpe, R. J., Bruce, M. A. & Norris, K. C. Poverty, Racism, and the Public Health Crisis in America. Front. Public Health 9, 699049 (2021).

25. Tannenbaum, S. L., Koru-Sengul, T., Zhao, W., Miao, F. & Byrne, M. M. Survival Disparities in Non– Small Cell Lung Cancer by Race, Ethnicity, and Socioeconomic Status. Cancer J. 20, 237–245 (2014).

26. Larsen, K., Rydz, E. & Peters, C. E. Inequalities in Environmental Cancer Risk and Carcinogen Exposures: A Scoping Review. Int. J. Environ. Res. Public. Health 20, 5718 (2023).

27. 2022 National Healthcare Quality and Disparities Report. (Agency for Healthcare Research and Quality (US), Rockville (MD), 2022).

28. Green, A. K. et al. Racial disparities in chemotherapy administration for early-stage breast cancer: a systematic review and meta-analysis. Breast Cancer Res. Treat. 172, 247–263 (2018).

29. Bakkila, B. F. et al. Evaluation of Racial Disparities in Quality of Care for Patients With Gastrointestinal Tract Cancer Treated With Surgery. *JAMA Netw*. Open 5, e225664 (2022).

30. Zhang, W., Edwards, A., Flemington, E. K. & Zhang, K. Racial disparities in patient survival and tumor mutation burden, and the association between tumor mutation burden and cancer incidence rate. Sci. Rep. 7, 13639 (2017).

31. Nassar, A. H., Adib, E. & Kwiatkowski, D. J. Distribution of *KRAS* ^G12C^ Somatic Mutations across Race, Sex, and Cancer Type. N. Engl. J. Med. 384, 185–187 (2021).

32. Bollig-Fischer, A. et al. Racial Diversity of Actionable Mutations in Non–Small Cell Lung Cancer. J. Thorac. Oncol. 10, 250–255 (2015).

33. Cote, M. L. et al. Racial Differences in Oncogene Mutations Detected in Early-Stage Low-Grade Endometrial Cancers. Int. J. Gynecol. Cancer 22, 1367–1372 (2012).

34. Arora, K. et al. Genetic Ancestry Correlates with Somatic Differences in a Real-World Clinical Cancer Sequencing Cohort. Cancer Discov. 12, 2552–2565 (2022).

35. Yuan, J. et al. Integrated Analysis of Genetic Ancestry and Genomic Alterations across Cancers. Cancer Cell 34, 549–560.e9 (2018).

36. Guttery, D. S. et al. Racial differences in endometrial cancer molecular portraits in The Cancer Genome Atlas. Oncotarget 9, 17093–17103 (2018).

37. Hebert-Magee, S. et al. The combined survival effect of codon 72 polymorphisms and p53 somatic mutations in breast cancer depends on race and molecular subtype. PLOS ONE 14, e0211734 (2019).

38. Ademuyiwa, F. O., Tao, Y., Luo, J., Weilbaecher, K. & Ma, C. X. Differences in the mutational landscape of triple-negative breast cancer in African Americans and Caucasians. Breast Cancer Res. Treat. 161, 491–499 (2017).

39. Munro, A. J., Lain, S. & Lane, D. P. P53 abnormalities and outcomes in colorectal cancer: a systematic review. Br. J. Cancer 92, 434–444 (2005).

40. Keenan, T. et al. Comparison of the Genomic Landscape Between Primary Breast Cancer in African American Versus White Women and the Association of Racial Differences With Tumor Recurrence. J. Clin. Oncol. Off. J. Am. Soc. Clin. Oncol. 33, 3621–3627 (2015).

41. Ansari-Pour, N. et al. Whole-genome analysis of Nigerian patients with breast cancer reveals ethnic-driven somatic evolution and distinct genomic subtypes. Nat. Commun. 12, 6946 (2021).

42. Bauml, J. et al. Frequency of EGFR and KRAS mutations in patients with non small cell lung cancer by racial background: Do disparities exist? Lung Cancer 81, 347–353 (2013).

43. Beroukhim, R. et al. The landscape of somatic copy-number alteration across human cancers. Nature 463, 899–905 (2010).

44. Taylor, A. M. et al. Genomic and Functional Approaches to Understanding Cancer Aneuploidy. Cancer Cell 33, 676–689.e3 (2018).

45. Lukow, D. A. & Sheltzer, J. M. Chromosomal instability and aneuploidy as causes of cancer drug resistance. Trends Cancer 8, 43–53 (2022).

46. Yang, S. Y. C., Pugh, T. J. & Oza, A. M. Double Trouble: Whole-Genome Doubling Distinguishes Early from Late Ovarian Cancer. Clin. Cancer Res. 28, 2730–2732 (2022).

47. Carrot-Zhang, J. et al. Comprehensive Analysis of Genetic Ancestry and Its Molecular Correlates in Cancer. Cancer Cell 37, 639–654.e6 (2020).

48. Levine, M. S. & Holland, A. J. The impact of mitotic errors on cell proliferation and tumorigenesis. Genes Dev. 32, 620–638 (2018).

49. Marei, H. E. et al. p53 signaling in cancer progression and therapy. Cancer Cell Int. 21, 703 (2021).

50. Vineis, P. & Husgafvel-Pursiainen, K. Air pollution and cancer: biomarker studies in human populations †. Carcinogenesis 26, 1846–1855 (2005).

51. Yu, X.-J. et al. Characterization of Somatic Mutations in Air Pollution-Related Lung Cancer. eBioMedicine 2, 583–590 (2015).

52. Bielski, C. M. et al. Genome doubling shapes the evolution and prognosis of advanced cancers. Nat. Genet. 50, 1189–1195 (2018).

53. Zeng, J., Hills, S. A., Ozono, E. & Diffley, J. F. X. Cyclin E-induced replicative stress drives p53-dependent whole-genome duplication. Cell 186, 528–542.e14 (2023).

54. Van de Peer, Y., Mizrachi, E. & Marchal, K. The evolutionary significance of polyploidy. Nat. Rev. Genet. 18, 411–424 (2017).

55. Frankell, A. M. et al. The evolution of lung cancer and impact of subclonal selection in TRACERx. Nature 616, 525–533 (2023).

56. Prasad, K. et al. Whole-Genome Duplication Shapes the Aneuploidy Landscape of Human Cancers. Cancer Res. 82, 1736–1752 (2022).

57. Lambuta, R. A. et al. Whole-genome doubling drives oncogenic loss of chromatin segregation. Nature 1–9 (2023) doi:10.1038/s41586-023-05794-2.

58. Nguyen, B. et al. Genomic characterization of metastatic patterns from prospective clinical sequencing of 25,000 patients. Cell 185, 563–575.e11 (2022).

59. The Cancer Genome Atlas homepage; http://cancergenome.nih.gov/abouttcga.

60. The ICGC/TCGA Pan-Cancer Analysis of Whole Genomes Consortium et al. Pan-cancer analysis of whole genomes. Nature 578, 82–93 (2020).

61. Spratt, D. E. et al. Racial/Ethnic Disparities in Genomic Sequencing. JAMA Oncol. 2, 1070–1074 (2016).

62. Landry, L. G., Ali, N., Williams, D. R., Rehm, H. L. & Bonham, V. L. Lack Of Diversity In Genomic Databases Is A Barrier To Translating Precision Medicine Research Into Practice. Health Aff. (Millwood) 37, 780–785 (2018).

63. Committee on the Use of Race, Ethnicity, and Ancestry as Population Descriptors in Genomics Research et al. Using Population Descriptors in Genetics and Genomics Research: A New Framework for an Evolving Field. 26902 (National Academies Press, Washington, D.C., 2023). doi:10.17226/26902.

64. Why Nature is updating its advice to authors on reporting race or ethnicity. Nature 616, 219–219 (2023).

65. Ashing, K. T., Jones, V., Bedell, F., Phillips, T. & Erhunmwunsee, L. Calling Attention to the Role of Race-Driven Societal Determinants of Health on Aggressive Tumor Biology: A Focus on Black Americans. JCO Oncol. Pract. 18, 15–22 (2022).

66. Shukla, A. et al. Chromosome arm aneuploidies shape tumour evolution and drug response. Nat. Commun. 11, 449 (2020).

67. Andreassen, P. R., Lohez, O. D., Lacroix, F. B. & Margolis, R. L. Tetraploid state induces p53-dependent arrest of nontransformed mammalian cells in G1. Mol. Biol. Cell 12, 1315–1328 (2001).

68. Ganem, N. J. et al. Cytokinesis Failure Triggers Hippo Tumor Suppressor Pathway Activation. Cell 158, 833–848 (2014).

69. López, S. et al. Interplay between whole-genome doubling and the accumulation of deleterious alterations in cancer evolution. Nat. Genet. 52, 283–293 (2020).

70. Quinton, R. J. et al. Whole-genome doubling confers unique genetic vulnerabilities on tumour cells. Nature 590, 492–497 (2021).

71. Dewhurst, S. M. et al. Tolerance of Whole-Genome Doubling Propagates Chromosomal Instability and Accelerates Cancer Genome Evolution. Cancer Discov. 4, 175–185 (2014).

72. Chang, E. & An, J.-Y. Whole-genome doubling is a double-edged sword: the heterogeneous role of whole-genome doubling in various cancer types. BMB Rep. 57, 125–134 (2024).

73. Kikutake, C. & Suyama, M. Pan-cancer analysis of whole-genome doubling and its association with patient prognosis. BMC Cancer 23, 619 (2023).

74. Lillard, J. W., Moses, K. A., Mahal, B. A. & George, D. J. Racial disparities in Black men with prostate cancer: A literature review. Cancer 128, 3787–3795 (2022).

75. Mahal, B. A. et al. Prostate Cancer Racial Disparities: A Systematic Review by the Prostate Cancer Foundation Panel. *Eur*. Urol. Oncol. 5, 18–29 (2022).

76. Hinata, N. & Fujisawa, M. Racial Differences in Prostate Cancer Characteristics and Cancer-Specific Mortality: An Overview. World J. Mens Health 40, 217–227 (2022).

77. Yamoah, K. et al. Racial and Ethnic Disparities in Prostate Cancer Outcomes in the Veterans Affairs Health Care System. *JAMA Netw*. Open 5, e2144027 (2022).

78. Li, K., Luo, H., Huang, L., Luo, H. & Zhu, X. Microsatellite instability: a review of what the oncologist should know. Cancer Cell Int. 20, 16 (2020).

79. Yamamoto, H. & Imai, K. Microsatellite instability: an update. Arch. Toxicol. 89, 899–921 (2015).

80. Popat, S., Hubner, R. & Houlston, R. S. Systematic review of microsatellite instability and colorectal cancer prognosis. J. Clin. Oncol. Off. J. Am. Soc. Clin. Oncol. 23, 609–618 (2005).

81. Zhu, L. et al. Microsatellite instability and survival in gastric cancer: A systematic review and meta-analysis. Mol. Clin. Oncol. 3, 699–705 (2015).

82. Madrigal, J. M. et al. Sociodemographic inequities in the burden of carcinogenic industrial air emissions in the United States. J. Natl. Cancer Inst. 116, 737–744 (2024).

83. Alvarez, C. H. Structural Racism as an Environmental Justice Issue: A Multilevel Analysis of the State Racism Index and Environmental Health Risk from Air Toxics. J. Racial Ethn. Health Disparities 10, 244– 258 (2023).

84. Bonner, S. N. et al. Structural Racism and Lung Cancer Risk: A Scoping Review. JAMA Oncol. 10, 122–128 (2024).

85. Rothstein, R. The Color of Law: A Forgotten History of How Our Government Segregated America. (Liveright Publishing, 2017).

86. Lane, H. M., Morello-Frosch, R., Marshall, J. D. & Apte, J. S. Historical Redlining Is Associated with Present-Day Air Pollution Disparities in U.S. Cities. Environ. Sci. Technol. Lett. 9, 345–350 (2022).

87. Zhang, L. et al. Association of Residential Racial and Economic Segregation With Cancer Mortality in the US. JAMA Oncol. 9, 122–126 (2023).

88. Brazil, N. Environmental inequality in the neighborhood networks of urban mobility in US cities. Proc. Natl. Acad. Sci. 119, e2117776119 (2022).

89. Kokot, H. et al. Prediction of Chronic Inflammation for Inhaled Particles: the Impact of Material Cycling and Quarantining in the Lung Epithelium. Adv. Mater. 32, 2003913 (2020).

90. Azawi, S. et al. Molecular cytogenetic characterization of the urethane-induced murine lung cell line LA-4 as a model for human squamous cell lung cancer. Mol. Clin. Oncol. 16, 9 (2022).

91. Watson, A. Y. & Valberg, P. A. Carbon black and soot: two different substances. AIHAJ J. Sci. Occup. Environ. Health Saf. 62, 218–228 (2001).

92. Jacobson, R. S., Korte, A. R., Vertes, A. & Miller, J. H. The Molecular Composition of Soot. Angew. Chem. Int. Ed Engl. 59, 4484–4490 (2020).

93. Jbaily, A. et al. Air pollution exposure disparities across US population and income groups. Nature 601, 228–233 (2022).

94. Mohai, P., Lantz, P. M., Morenoff, J., House, J. S. & Mero, R. P. Racial and Socioeconomic Disparities in Residential Proximity to Polluting Industrial Facilities: Evidence From the Americans’ Changing Lives Study. Am. J. Public Health 99, S649–S656 (2009).

95. Morello-Frosch, R. & Lopez, R. The riskscape and the color line: examining the role of segregation in environmental health disparities. Environ. Res. 102, 181–196 (2006).

96. Kucab, J. E. et al. A Compendium of Mutational Signatures of Environmental Agents. Cell 177, 821–836.e16 (2019).

97. Abdel-Shafy, H. I. & Mansour, M. S. M. A review on polycyclic aromatic hydrocarbons: Source, environmental impact, effect on human health and remediation. Egypt. J. Pet. 25, 107–123 (2016).

98. Kyjovska, Z. O. et al. DNA damage following pulmonary exposure by instillation to low doses of carbon black (Printex 90) nanoparticles in mice. Environ. Mol. Mutagen. 56, 41–49 (2015).

99. Boffetta, P., Jourenkova, N. & Gustavsson, P. Cancer risk from occupational and environmental exposure to polycyclic aromatic hydrocarbons. Cancer Causes Control CCC 8, 444–472 (1997).

100. Corrêa, S. M., Arbilla, G., da Silva, C. M., Martins, E. M. & de Souza, S. L. Q. Determination of carbonyls and size-segregated polycyclic aromatic hydrocarbons, and their nitro and alkyl analogs in emissions from diesel–biodiesel-ethanol blends. Environ. Sci. Pollut. Res. 30, 62470–62480 (2023).

101. Garg, S. Towards routine chromosome-scale haplotype-resolved reconstruction in cancer genomics. Nat. Commun. 14, 1358 (2023).

102. Jaratlerdsiri, W. et al. African-specific molecular taxonomy of prostate cancer. Nature 609, 552–559 (2022).

103. Gong, T. et al. Genome-wide interrogation of structural variation reveals novel African-specific prostate cancer oncogenic drivers. Genome Med. 14, 100 (2022).

104. Tessum, C. W. et al. PM _2.5_ polluters disproportionately and systemically affect people of color in the United States. Sci. Adv. 7, eabf4491 (2021).

105. Hieronymus, H. et al. Tumor copy number alteration burden is a pan-cancer prognostic factor associated with recurrence and death. eLife 7, e37294 (2018).

106. Gómez-Rueda, H., Martínez-Ledesma, E., Martínez-Torteya, A., Palacios-Corona, R. & Trevino, V. Integration and comparison of different genomic data for outcome prediction in cancer. BioData Min. 8, 32 (2015).

107. Van Dijk, E. et al. Chromosomal copy number heterogeneity predicts survival rates across cancers. Nat. Commun. 12, 3188 (2021).

108. Smith, J. C. & Sheltzer, J. M. Systematic identification of mutations and copy number alterations associated with cancer patient prognosis. eLife 7, e39217 (2018).

109. Girish, V. et al. Oncogene-like addiction to aneuploidy in human cancers. Science 381, eadg4521 (2023).

110. Cohen-Sharir, Y. et al. Aneuploidy renders cancer cells vulnerable to mitotic checkpoint inhibition. Nature 590, 486–491 (2021).

111. Govindan, R. et al. Trial in progress: A phase 1, multicenter, open-label, dose-exploration and dose-expansion study evaluating the safety, tolerability, pharmacokinetics, and efficacy of AMG650 in subjects with advanced solid tumors. J. Clin. Oncol. 39, TPS5600–TPS5600 (2021).

112. Cerami, E. et al. The cBio Cancer Genomics Portal: An Open Platform for Exploring Multidimensional Cancer Genomics Data. Cancer Discov. 2, 401–404 (2012).

113. Gao, J. et al. Integrative Analysis of Complex Cancer Genomics and Clinical Profiles Using the cBioPortal. Sci. Signal. 6, (2013).

114. Shen, R. & Seshan, V. E. FACETS: allele-specific copy number and clonal heterogeneity analysis tool for high-throughput DNA sequencing. Nucleic Acids Res. 44, e131–e131 (2016).

115. Carter, S. L. et al. Absolute quantification of somatic DNA alterations in human cancer. Nat. Biotechnol. 30, 413–421 (2012).

116. Nik-Zainal, S. A compendium of mutational signatures of environmental agents Kucab, et al. Mendeley 10.17632/M7R4MSJB4C.2 (2019).

117. R Core Team. R: A Language and Environment for Statistical Computing. https://www.R-project.org/ (2022).

118. Van Rossum, G. & Drake, F. L. Python 3 Reference Manual. (CreateSpace, Scotts Valley, CA, 2009).

119. Fox, J. & Weisberg, S. An R Companion to Applied Regression. (SAGE, Los Angeles London New Delhi Singapore Washington, DC Melbourne, 2019).

120. Blighe, K. EnhancedVolcano. Bioconductor 10.18129/B9.BIOC.ENHANCEDVOLCANO (2018).

121. Wickham, H. Ggplot2: Elegant Graphics for Data Analysis. (Springer International Publishing : Imprint: Springer, Cham, 2016). doi:10.1007/978-3-319-24277-4.

122. Iannone, R., et al. Gt: Easily Create Presentation-Ready Display Tables. https://CRAN.R-project.org/package=gt (2023).

123. Sjoberg, D., D., Whiting, K., Curry, M., Lavery, J., A. & Larmarange, J. Reproducible Summary Tables with the gtsummary Package. R J. 13, 570 (2021).

124. Mayakonda, A., Lin, D.-C., Assenov, Y., Plass, C. & Koeffler, H. P. Maftools: efficient and comprehensive analysis of somatic variants in cancer. Genome Res. 28, 1747–1756 (2018).

125. Schauberger, P. & Walker, A. Openxlsx: Read, Write and Edit Xlsx Files. https://CRAN.R-project.org/package=openxlsx (2023).

126. Wickham, H. Reshaping Data with the reshape Package. J. Stat. Softw. 21, (2007).

127. Wickham, H. et al. Welcome to the tidyverse. J. Open Source Softw. 4, 1686 (2019).

128. Davidson-Pilon, C. lifelines: survival analysis in Python. J. Open Source Softw. 4, 1317 (2019).

129. Hunter, J. D. Matplotlib: A 2D Graphics Environment. Comput. Sci. Eng. 9, 90–95 (2007).

130. Team, T. P. D. pandas-dev/pandas: Pandas. Zenodo 10.5281/ZENODO.3509134 (2023).

131. Abberior Instruments Development Team. Imspector Image Acquisition & Analysis Software v16.2.

132. ImageJ2. ImageJ Wiki https://imagej.github.io/software/imagej2/index.

133. Wolfram Research, Inc. Mathematica, Version 12.0. (2019).

134. Degasperi, A. et al. A practical framework and online tool for mutational signature analyses show intertissue variation and driver dependencies. Nat. Cancer 1, 249–263 (2020).

135. Benjamini, Y. & Hochberg, Y. Controlling the False Discovery Rate: A Practical and Powerful Approach to Multiple Testing. J. R. Stat. Soc. Ser. B Methodol. 57, 289–300 (1995).

136. National Institute of Standards and Technology. Certificate of Analysis Standard Reference Material 1650b. (2021).

137. Bendtsen, K. M. et al. Particle characterization and toxicity in C57BL/6 mice following instillation of five different diesel exhaust particles designed to differ in physicochemical properties. Part. Fibre Toxicol. 17, 38 (2020).

138. Bourdon, J. A. et al. Carbon black nanoparticle instillation induces sustained inflammation and genotoxicity in mouse lung and liver. Part. Fibre Toxicol. 9, 5 (2012).

139. Karl, A. et al. Aqueous carbon black dispersions.

140. Di Ianni, E. et al. Assessment of primary and inflammation-driven genotoxicity of carbon black nanoparticles in vitro and in vivo. Nanotoxicology 16, 526–546 (2022).

141. Rouzaud, J.-N., Duber, S., Pawlyta, M., Cacciaguerra, T. & Clinard, C. TEM study of carbon nanoparticles. Relationships multiscale organization - properties. Proc. Am. Carbon Soc. (2004).

142. Bengtson, S. et al. No cytotoxicity or genotoxicity of graphene and graphene oxide in murine lung epithelial FE1 cells in vitro. Environ. Mol. Mutagen. 57, 469–482 (2016).

143. Rasmussen, K. et al. Multi-walled Carbon Nanotubes, NM-400, NM-401, NM-402, NM-403: Characterisation and Physico-Chemical Properties. JRC Publications Repository https://publications.jrc.ec.europa.eu/repository/handle/JRC91205 (2014) doi:10.2788/10986.

144. Bornholdt, J. et al. Identification of Gene Transcription Start Sites and Enhancers Responding to Pulmonary Carbon Nanotube Exposure in Vivo. ACS Nano 11, 3597–3613 (2017).

145. Barfod, K. K. et al. Increased surface area of halloysite nanotubes due to surface modification predicts lung inflammation and acute phase response after pulmonary exposure in mice. Environ. Toxicol. Pharmacol. 73, 103266 (2020).

146. Di Ianni, E. et al. Organomodified nanoclays induce less inflammation, acute phase response, and genotoxicity than pristine nanoclays in mice lungs. Nanotoxicology 14, 869–892 (2020).

147. Hadrup, N. et al. Pulmonary toxicity of Fe2O3, ZnFe2O4, NiFe2O4 and NiZnFe4O8 nanomaterials: Inflammation and DNA strand breaks. Environ. Toxicol. Pharmacol. 74, 103303 (2020).

148. Cho, W.-S. et al. Inflammatory mediators induced by intratracheal instillation of ultrafine amorphous silica particles. Toxicol. Lett. 175, 24–33 (2007).

149. Umek, P., Korošec, R. C., Jančar, B., Dominko, R. & Arčon, D. The Influence of the Reaction Temperature on the Morphology of Sodium Titanate 1D Nanostructures and Their Thermal Stability. J. Nanosci. Nanotechnol. 7, 3502–3508 (2007).

